# Evaluation of Domain Generalization and Adaptation on Improving Model Robustness to Temporal Dataset Shift in Clinical Medicine

**DOI:** 10.1101/2021.06.17.21259092

**Authors:** Lin Lawrence Guo, Stephen R Pfohl, Jason Fries, Alistair Johnson, Jose Posada, Catherine Aftandilian, Nigam Shah, Lillian Sung

## Abstract

**Importance:** Temporal dataset shift associated with changes in healthcare over time is a barrier to deploying machine learning-based clinical decision support systems. Algorithms that learn robust models by estimating invariant properties across time periods for domain generalization (DG) and unsupervised domain adaptation (UDA) might be suitable to proactively mitigate dataset shift.

**Objective:** To characterize the impact of temporal dataset shift on clinical prediction models and benchmark DG and UDA algorithms on improving model robustness.

**Design, Setting, and Participants:** In this cohort study, intensive care unit patients from the MIMIC-IV database were categorized by year groups (2008–2010, 2011–2013, 2014–2016 and 2017–2019). Tasks were predicting mortality, long length of stay, sepsis and invasive ventilation. Feedforward neural networks were used as prediction models. The baseline experiment trained models using empirical risk minimization (ERM) on 2008–2010 (ERM[08-10]) and evaluated them on subsequent year groups. DG experiment trained models using algorithms that estimated invariant properties using 2008–2016 and evaluated them on 2017– 2019. UDA experiment leveraged unlabelled samples from 2017–2019 for unsupervised distribution matching. DG and UDA models were compared to ERM[08-16] models trained using 2008-2016.

**Main Outcome(s) and Measure(s):** Main performance measures were area-under-the-receiver-operating-characteristic curve (AUROC), area-under-the-precision-recall curve and absolute calibration error. Threshold-based metrics including false-positives and false-negatives were used to assess the clinical impact of temporal dataset shift and its mitigation strategies.

**Results:** In the baseline experiments, dataset shift was most evident for sepsis prediction (maximum AUROC drop, 0.090; 95% confidence interval (CI), 0.080-0.101). Considering a scenario of 100 consecutively admitted patients showed that ERM[08-10] applied to 2017-2019 was associated with one additional false-negative among 11 patients with sepsis, when compared to the model applied to 2008-2010. When compared with ERM[08-16], DG and UDA experiments failed to produce more robust models (range of AUROC difference, −0.003-0.050).

**Conclusions and Relevance:** DG and UDA failed to produce more robust models compared to ERM in the setting of temporal dataset shift. Alternate approaches are required to preserve model performance over time in clinical medicine.

**KEY POINTS:** *Question:* Can algorithms that estimate invariant properties across environments for domain generalization and unsupervised domain adaptation improve the robustness of machine learning-derived clinical prediction models to temporal dataset shift?

*Findings:* In this cohort study using 4 clinical outcomes, domain generalization and unsupervised domain adaptation algorithms did not meaningfully outperform the standard model training algorithm – empirical risk minimization – in learning robust models that generalize over time in the presence of temporal dataset shift.

*Meaning:* These findings highlight the difficulty of improving robustness to dataset shift with purely data-driven techniques that do not leverage prior knowledge of the nature of the shift and the requirement of alternate approaches to preserve model performance over time in clinical medicine.

## INTRODUCTION

The wide-spread adoption of electronic health records (EHRs) and the enhanced capacity to store and perform computation with large amounts of data have enabled the development of highly performant machine learning models for clinical outcome predictions.^1^ The utility of these models critically depends on sustained performance to maintain safety,^2^ end-users’ trust, and to outweigh the high cost of integrating each model into the clinical workflow.^3^ However, this is hindered in the non-stationary healthcare environment by temporal dataset shift due to mismatch between the data distribution on which models were developed and the distribution to which models were applied.^4^

There has been limited research on the impact of temporal dataset shift in clinical medicine.^5^ Recent approaches largely relied on maintenance strategies consisting of performance monitoring, model updating and calibration over certain time intervals.^6-8^ Another approach grouped clinical features into their underlying concepts to cope with a change in the record-keeping system.^9^ Generally, these approaches either require detection of model degradation or rely on explicit knowledge or assumptions about the underlying cause of the shift. Complementary to these approaches would be ones that attempt to proactively produce robust models that incorporate relatively few assumptions on the nature of the shift.

The past decade of machine learning research offered numerous algorithms that learn robust models by using data from multiple environments to identify invariant properties. These algorithms were often developed for domain generalization (DG)^10^ and unsupervised domain adaptation (UDA).^11^ In the DG setting, the goal is to learn models that generalize to new environments unseen at training time. In the UDA setting, the goal is to adapt models to target environments using labeled samples from the source environment as well as a limited set of unlabeled samples from the target environment. If we consider EHR data across discrete time windows as related but distinct environments, then both DG and UDA settings may be suitable to combat the impact of temporal dataset shift. To date, these approaches have not been evaluated on improving model robustness to temporal dataset shift for clinical prediction tasks. Therefore, the objective was to benchmark learning algorithms for DG and UDA on mitigating the impact of temporal dataset shift on machine learning model performance in a set of clinical prediction tasks.

## METHODS

### Data Source

We used the MIMIC-IV database,^12^ which contains deidentified EHRs of 382,278 patients admitted to an intensive care unit (ICU) or the emergency department at the Beth Israel Deaconess Medical Center (BIDMC) between 2008–2019. For this cohort study, we considered ICU admissions sourced from the clinical information system MetaVision at the BIDMC, in which records from 53,150 patients were made available in the latest version of MIMIC-IV 1.0. Because of deidentification, the requirement for Institutional Review Board approval was waived.

### Cohort

Each patient’s timeline in MIMIC-IV is anchored to a shifted (deidentified) year with which a *year group* and *age* are associated. The year group reflects the actual 3-year range (for example 2008–2010) in which the shifted year occurred, and age reflects the patient’s actual age in the shifted year. There are four available year groups in MIMIC-IV: 2008–2010, 2011–2013, 2014–2016 and 2017–2019. We included patients who were 18 years or older and randomly selected one ICU admission that occurred in the year group for each patient. As a result, each patient is represented once in our dataset and is associated with a single year group. We excluded ICU admissions less than 4 hours in duration.

### Outcomes

We defined four clinical outcomes. For each outcome, the task was to perform binary predictions over a time horizon with respect to the time of prediction, which was set as 4 hours after ICU admission. Long length of stay (*Long LOS*) was defined as ICU stay greater than three days from the prediction time. *Mortality* corresponded to in-hospital mortality within 7 days from the prediction time. *Invasive ventilation* corresponded to initiation of invasive ventilation within 24 hours from the prediction time. *Sepsis* corresponded to the development of sepsis according to the Sepsis-3 criteria^13^ within 7 days from the prediction time. For invasive ventilation and sepsis, we excluded patients with these outcomes prior to the time of prediction. Further details on each outcome are presented in the **eMethods** in the Supplement.

### Features

Our feature extraction followed a common procedure^14^ and obtained six categories of features including diagnoses, procedures, labs, prescriptions, ICU charts and demographics. Demographic features included age, biological sex, race, insurance, marital status and language. Clinical features were extracted over a set of time-intervals defined relative to the time of ICU admission as follows: 0-4 hours after ICU admission, 0-7 days prior, 7-30 days prior, 30-180 days prior, and 180 days-any time prior. For each time interval, we obtained counts of unique concept identifiers for diagnoses, procedures, prescriptions and labs with the exception that identifiers for diagnoses and procedures were not obtained in the 0-4 hours interval after admission as they were not available. We also obtained measurements for lab tests for each time interval, and measurements for chart events in the 0-4 hours interval after admission. In addition, we mapped each measurement variable in each time interval to the patient-level mean, minimum and maximum. Number of extracted features for each category and time interval are listed in the **eMethods** in the Supplement.

Feature preprocessing pruned features that had less than 25 patient observations, replaced non-zero count values with 1s, encoded measurement features to quintiles, and one-hot encoded all but count features. This process resulted in binary feature matrices that were extremely sparse. All feature preprocessing procedures were fit on the training set (e.g., to determine the boundaries of each quintile) and were subsequently applied to transform the validation and test sets.

### Model and Learning Algorithms

In all experiments, we leveraged fully connected feedforward neural network (NN) models for prediction, as they enable flexible learning for differentiable objectives across algorithms. The standard algorithm to learn models is the *empirical risk minimization* (ERM) algorithm in which the objective is to minimize average training error^15^ without considerations of environment annotations (year groups in our case).

Algorithms for DG and UDA differ on the types of invariance they assume. *Invariant risk minimization* (IRM)^16^ is a DG algorithm that learns a latent representation (i.e., hidden layer activations) where the optimal classifier leveraging that representation is the same for all environments. *Group distributionally robust optimization* (GroupDRO)^17^ is a DG algorithm that does not “learn” invariances but instead minimizes training error in the worst-case training environment by increasing the importance of environments with larger errors. Algorithms that learn to match latent representation across environments can be leveraged for DG as well as UDA since these algorithms do not require outcome labels. These include *correlation alignment* (CORAL),^18^ which seeks to match the mean and covariance of the distribution of the data encoded in the latent space across environments, and *domain adversarial learning* (AL),^19,20^ which matches the distributions using an adversarial network and an objective that minimizes discriminability between environments. The adversarial network used in this study is a NN model with one hidden layer of dimension 32.

### Model Development

We conducted baseline, DG and UDA experiments. The *baseline* experiment consisted of several aspects. First, to characterize temporal dataset shift on model performance, we trained models with ERM on the 2008–2010 group (ERM[08-10]) and evaluated these models in each subsequent year group.

Next, to describe the extent of temporal dataset shift, we compared the performance of ERM[08-10] in each subsequent year group with models trained using ERM on that year group. Difference in performance in the target year group (2017-2019) between ERM[08-10] and models trained and evaluated on the target year group (ERM[17-19]) described the extent of temporal dataset shift in the extreme scenario in which models were developed on the earliest available data and were never updated. All models in the baseline experiment used ERM.

For *DG* and *UDA* experiments, model training was performed on 2008–2016, with UDA also incorporating unlabelled samples from the target year group. Performance of DG and UDA models were compared with ERM models trained using 2008-2016 (ERM[08-16]) as these are the fairest ERM comparators for DG and UDA models. For models in the DG and UDA experiments, we focused on their performance in the target year group, but also described their performance in 2008-2016.

#### Data splitting procedure

Data splitting procedure was performed separately for each task and experiment (see **Figure 1**). The baseline experiment split each year group into 70% training, 15% validation and 15% test sets. In DG and UDA experiments, the training set included 85% data from 2008–2010 and 2011–2013, and 45% of data from 2014–2016. The validation set included 35% of data from 2014–2016 (chosen because of its temporal proximity to the target year group). The test set included the same 15% from each year group as the baseline experiment, which allowed us to compare model performance across experiments and learning algorithms on the same patients. For UDA, training year groups were combined into one group, and unlabeled samples of various sizes (100, 500, 1000, and 1500) from the target year group (2017–2019) were leveraged for unsupervised distribution matching.

**Figure 1.**
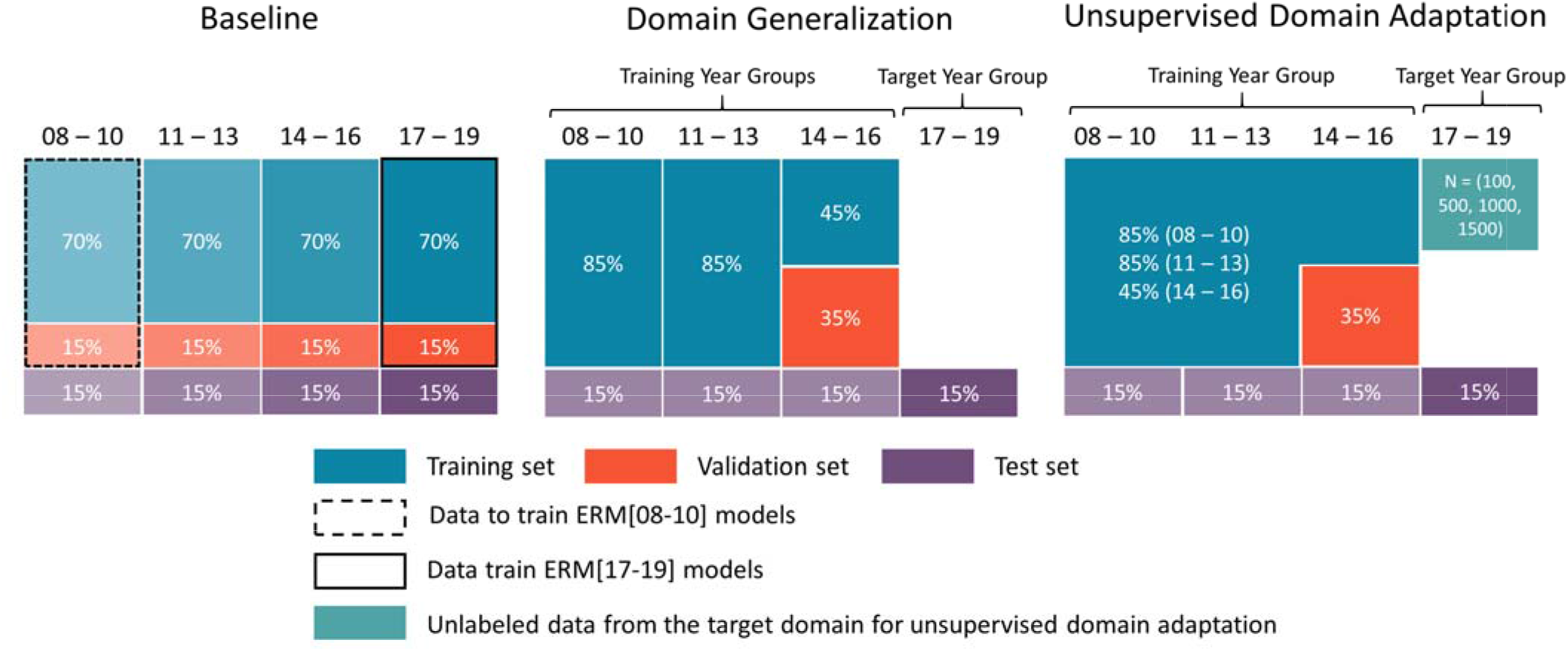
Data Splitting Procedure for baseline, domain generalization (DG) and unsupervised domain adaptation (UDA) experiments. Different shades of the same color indicate that they were used to train or evaluate different models. For instance, in the baseline experiment, the training set of each year group was used to learn models for that year group. In the DG experiment, the training year groups were kept separate to allow DG algorithms to estimate invariance across the year groups. In comparison, in the UDA experiment, data from the training year groups were pooled, and unlabeled samples from the target year group were leveraged for unsupervised distribution matching between training and target year groups. In addition, ERM[08-16] models were learned on pooled data from the training year groups (2008-2016) to be used as ERM comparators for DG and UDA models.

#### Model training

We developed NN models on the training sets of each experiment for each task and selected hyperparameters based on performance in the validation sets. DG and UDA models used the same model hyperparameters as ERM[08-16], but involved an additional search over the algorithm-specific hyperparameter that modulated the impact of the algorithm on model learning. For all experiments, we trained 20 NN models using the selected hyperparameters for each combination of outcome, learning algorithm and experiment-specific characteristic (for example, year group in baseline experiment and size of unlabeled samples for UDA). Further details on the models, learning algorithms, as well as hyperparameter selection and model training procedures are presented in the **eMethods** in the Supplement.

#### Model evaluation

We evaluated models on the test sets of each experiment. Model performance was evaluated using area-under-receiver-operating-characteristic curve (AUROC), area-under-precision-recall curve (AUPRC) and the absolute calibration error (ACE).^21^ ACE is a calibration measure similar to the integrated calibration index^22^ in that it assesses overall model calibration by taking the average of the absolute deviations from an approximated perfect calibration curve. The difference is that ACE uses logistic regression for approximation instead of locally weighted regression such as LOESS.

To aid clinical interpretation of the impact of temporal dataset shift and its mitigation strategies, we translated change in performance to interpretable threshold-based metrics (including sensitivity and specificity) across clinically reasonable threshold levels. We chose the task with the most extreme temporal dataset shift. We setup a scenario with 100 hypothetical ICU patients and estimated the number of patients with and without a positive label using average prevalence from 2018 to 2019. We then illustrated the number of false-positive-(FP) and false-negative predictions (FN) for: (1) ERM[08-10] in 2008-2010, illustrating the results of initial model development with training and test sets in 2008-2010, and representing performance anticipated by clinicians applying the model to patients admitted in 2017-2019 if the model is not updated; (2) ERM[08-10] in 2017-2019, illustrating the actual performance of the earlier model on patients, or the impact of temporal dataset shift; (3) ERM[08-16] in 2017-2019, illustrating the ERM comparators for DG and UDA models; (4) models trained using a representative approach from DG or UDA; and (5) ERM[17-19] in which training and test sets are both using 2017-2019 data.

### Statistical Analysis

For each combination of outcome, experiment-specific characteristic (e.g., year group in the baseline experiment), and evaluation metric (AUROC, AUPRC and ACE), we reported the median and 95% confidence interval (CI) of the distribution over mean performance (across 20 NN models) in the test set obtained from 10,000 bootstrap iterations. To compare models (for example, learned using IRM vs. ERM[08-16]) in the target year group, metrics were computed over 10,000 bootstrap iterations and the resulting 95% confidence interval of the differences were used to determine statistical significance.^23^

Model training was performed on an Nvidia V100 GPU. Analyses were implemented in Python 3.8,^24^ Scikit-learn 0.24^25^ and Pytorch 1.7^26^. The code for all analyses is open-source and available online^a^.

## RESULTS

Cohort characteristics for each year group and outcome are presented in **Table 1. Figure 2** shows performance measures (AUROC, AUPRC and ACE) of ERM[08-10] models in each year group vs. models trained on that year group. Largest temporal dataset shift was observed for sepsis predictions in 2017-2019 (drop in AUROC, 0.090; 95% CI, 0.080-0.101).

**Table 1.** Cohort Characteristics by Year Group.

|  | 2008 – 2010 | 2011 – 2013 | 2014 – 2016 | 2017 - 2019 |
| --- | --- | --- | --- | --- |
| **Mortality** |  |  |  |  |
| Patients, No. (% pos) | 9042 (7.4%) | 9476 (7.1%) | 10289 (7.4%) | 10060 (7.2%) |
| Age, mean ± SD | 63±18 | 63±18 | 64±17 | 64±17 |
| Sex, No. (%) |  |  |  |  |
| Female | 3864 (43%) | 4090 (43%) | 4430 (43%) | 4170 (41%) |
| Male | 5178 (57%) | 5386 (57%) | 5859 (57%) | 5890 (59%) |
| Race, No. (%) |  |  |  |  |
| White | 6784 (75%) | 6217 (66%) | 6468 (63%) | 6129 (61%) |
| Other | 2258 (25%) | 3259 (34%) | 3821 (37%) | 3931 (39%) |
| **Long Length of Stay** |  |  |  |  |
| Patients, No. (% pos) | 9042 (29.8%) | 9476 (28.4%) | 10289 (31.0%) | 10060 (35.2%) |
| Age, mean ± SD | 63±18 | 63±18 | 64±17 | 64±17 |
| Sex, No. (%) |  |  |  |  |
| Female | 3864 (43%) | 4090 (43%) | 4430 (43%) | 4170 (41%) |
| Male | 5178 (57%) | 5386 (57%) | 5859 (57%) | 5890 (59%) |
| Race, No. (%) |  |  |  |  |
| White | 6784 (75%) | 6217 (66%) | 6468 (63%) | 6129 (61%) |
| Other | 2258 (25%) | 3259 (34%) | 3821 (37%) | 3931 (39%) |
| **Invasive Ventilation** |  |  |  |  |
| Patients, No. (% pos) | 6692 (10.2%) | 7181 (10.1%) | 7447 (12.1%) | 7311 (11.4%) |
| Age, mean ± SD | 64±18 | 64±18 | 64±17 | 64±17 |
| Sex, No. (%) |  |  |  |  |
| Female | 2947 (44%) | 3133 (44%) | 3318 (45%) | 3124 (43%) |
| Male | 3745 (56%) | 4048 (56%) | 4129 (55%) | 4187 (57%) |
| Race, No. (%) |  |  |  |  |
| White | 5078 (76%) | 4839 (67%) | 4920 (66%) | 4727 (65%) |
| Other | 1614 (24%) | 2342 (33%) | 2527 (34%) | 2584 (35%) |
| **Sepsis** |  |  |  |  |
| Patients, No. (% pos) | 5410 (12.7%) | 5557 (10.2%) | 6217 (10.8%) | 7161 (10.6%) |
| Age, mean ± SD | 62±19 | 62±18 | 62±18 | 63±17 |
| Sex, No. (%) |  |  |  |  |
| Female | 2334 (43%) | 2442 (44%) | 2809 (45%) | 2948 (41%) |
| Male | 3076 (57%) | 3115 (56%) | 3408 (55%) | 4213 (59%) |
| Race, No. (%) |  |  |  |  |
| White | 4032 (75%) | 3648 (66%) | 3945 (63%) | 4447 (62%) |
| Other | 1378 (25%) | 1909 (34%) | 2272 (37%) | 2714 (38%) |

Abbreviations. pos: positive labels; SD: standard deviation.

**Figure 2.**
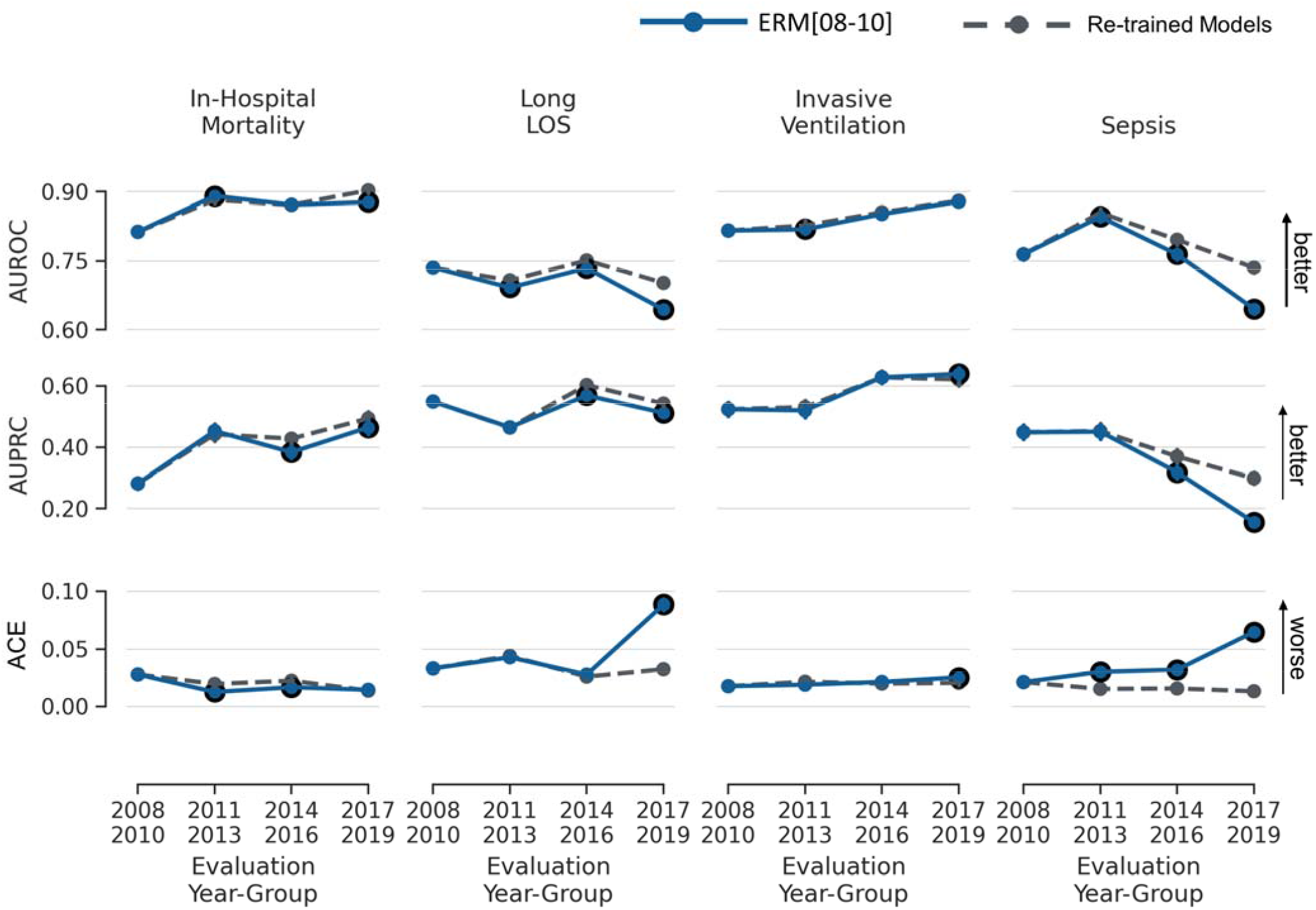
Mean performance (AUROC, AUPRC, and ACE) of models in the baseline experiment. Solid blue lines depict models trained using 2008-2010 (ERM[08-10]) and evaluated in each year group. Dashed lines depict models trained and evaluated in each year group separately (comparators). Error bars indicate 95% confidence interval obtained from 10,000 bootstrap iterations. Black circles indicate statistically significant differences in performance based on the 95% confidence interval of the difference over 10,000 bootstrap iterations when comparing ERM[08-10] and comparators for each year group. The figure shows temporal dataset shift that is larger for Long LOS and Sepsis tasks. Abbreviations, ER: empirical risk minimization; LOS: length of stay; AUROC: area under the receiver operating characteristics curve; AUPRC: area under the precision recall curve; ACE: absolute calibration error.

Figure 3 illustrates change in the performance measures of DG and UDA models in the target year group (2017 – 2019) relative to ERM[08-16]. In addition, change in performance measures of ERM[08-10] and ERM[17-19] are plotted in grey for comparison. ERM[08-16] performed better than ERM[08-10] (largest gain in AUROC, 0.049; 95% CI, 0.041-0.057; **eTable 1** in the Supplement), but performed worse than ERM[17-19] (worst drop in AUROC, 0.071; 95% CI, 0.062-0.081) with some exceptions in mortality and invasive ventilation predictions (**eTable 2** in the Supplement). Performance of DG and UDA models was similar to ERM[08-16] and while some models performed significantly better than ERM[08-16], others performed significantly worse with all differences being relatively small in magnitude (**eTable 3** in the Supplement). In addition, increasing the magnitude of the algorithm-specific hyperparameters of the DG and UDA algorithms did not result in performance gains (see **eFigures 1, 2**, and **3** in the Supplement).

**Table 2** illustrates a clinical interpretation of temporal dataset shift in sepsis prediction using a scenario of 100 consecutively admitted patients to the ICU between 2017-2019 with a risk threshold of 10% and an estimated outcome prevalence of 11% (see **eTable 5** in the Supplement for results across thresholds from 5% to 45%). ERM[08-10] applied to 2017-2019 was associated with one additional FN among 11 patients with sepsis and 7 additional FP among 89 patients without sepsis when compared to the model applied to 2008-2010. FN with AL, as the representative mitigation approach, was similar to ERM[08-16].

**Table 2.**
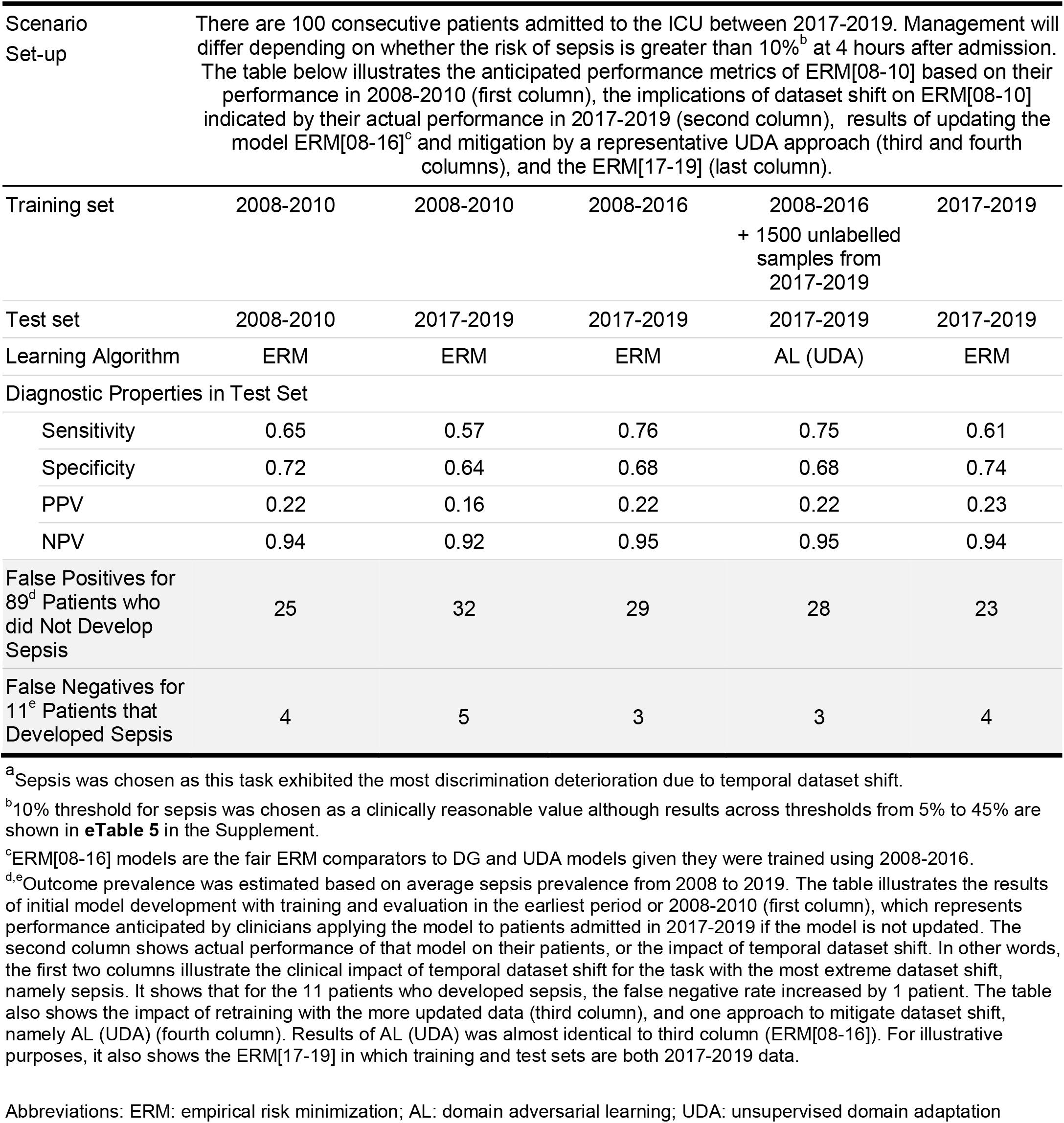
Clinical Interpretation of Temporal Dataset Shift in Sepsis Prediction^a^

|  |  |  |  |  |  |
| --- | --- | --- | --- | --- | --- |
| Scenario Set-up | There are 100 consecutive patients admitted to the ICU between 2017-2019. Management will differ depending on whether the risk of sepsis is greater than 10%<sup>b</sup> at 4 hours after admission. The table below illustrates the anticipated performance metrics of ERM[08-10] based on their performance in 2008-2010 (first column), the implications of dataset shift on ERM[08-10] indicated by their actual performance in 2017-2019 (second column), results of updating the model ERM[08-16]<sup>c</sup> and mitigation by a representative UDA approach (third and fourth columns), and the ERM[17-19] (last column). |  |  |  |  |
| Training set | 2008-2010 | 2008-2010 | 2008-2016 | 2008-2016<br>+ 1500 unlabelled samples from 2017-2019 | 2017-2019 |
| Test set | 2008-2010 | 2017-2019 | 2017-2019 | 2017-2019 | 2017-2019 |
| Learning Algorithm | ERM | ERM | ERM | AL (UDA) | ERM |
| Diagnostic Properties in Test Set |  |  |  |  |  |
| Sensitivity | 0.65 | 0.57 | 0.76 | 0.75 | 0.61 |
| Specificity | 0.72 | 0.64 | 0.68 | 0.68 | 0.74 |
| PPV | 0.22 | 0.16 | 0.22 | 0.22 | 0.23 |
| NPV | 0.94 | 0.92 | 0.95 | 0.95 | 0.94 |
| False Positives for 89<sup>d</sup> Patients who did Not Develop Sepsis | 25 | 32 | 29 | 28 | 23 |
| False Negatives for 11<sup>e</sup> Patients that Developed Sepsis | 4 | 5 | 3 | 3 | 4 |

<sup>a</sup>Sepsis was chosen as this task exhibited the most discrimination deterioration due to temporal dataset shift.
<sup>b</sup>10% threshold for sepsis was chosen as a clinically reasonable value although results across thresholds from 5% to 45% are shown in **eTable 5** in the Supplement.
<sup>c</sup>ERM[08-16] models are the fair ERM comparators to DG and UDA models given they were trained using 2008-2016.
<sup>d,e</sup>Outcome prevalence was estimated based on average sepsis prevalence from 2008 to 2019. The table illustrates the results of initial model development with training and evaluation in the earliest period or 2008-2010 (first column), which represents performance anticipated by clinicians applying the model to patients admitted in 2017-2019 if the model is not updated. The second column shows actual performance of that model on their patients, or the impact of temporal dataset shift. In other words, the first two columns illustrate the clinical impact of temporal dataset shift for the task with the most extreme dataset shift, namely sepsis. It shows that for the 11 patients who developed sepsis, the false negative rate increased by 1 patient. The table also shows the impact of retraining with the more updated data (third column), and one approach to mitigate dataset shift, namely AL (UDA) (fourth column). Results of AL (UDA) was almost identical to third column (ERM[08-16]). For illustrative purposes, it also shows the ERM[17-19] in which training and test sets are both 2017-2019 data.
Abbreviations: ERM: empirical risk minimization; AL: domain adversarial learning; UDA: unsupervised domain adaptation

**Figure 3.**
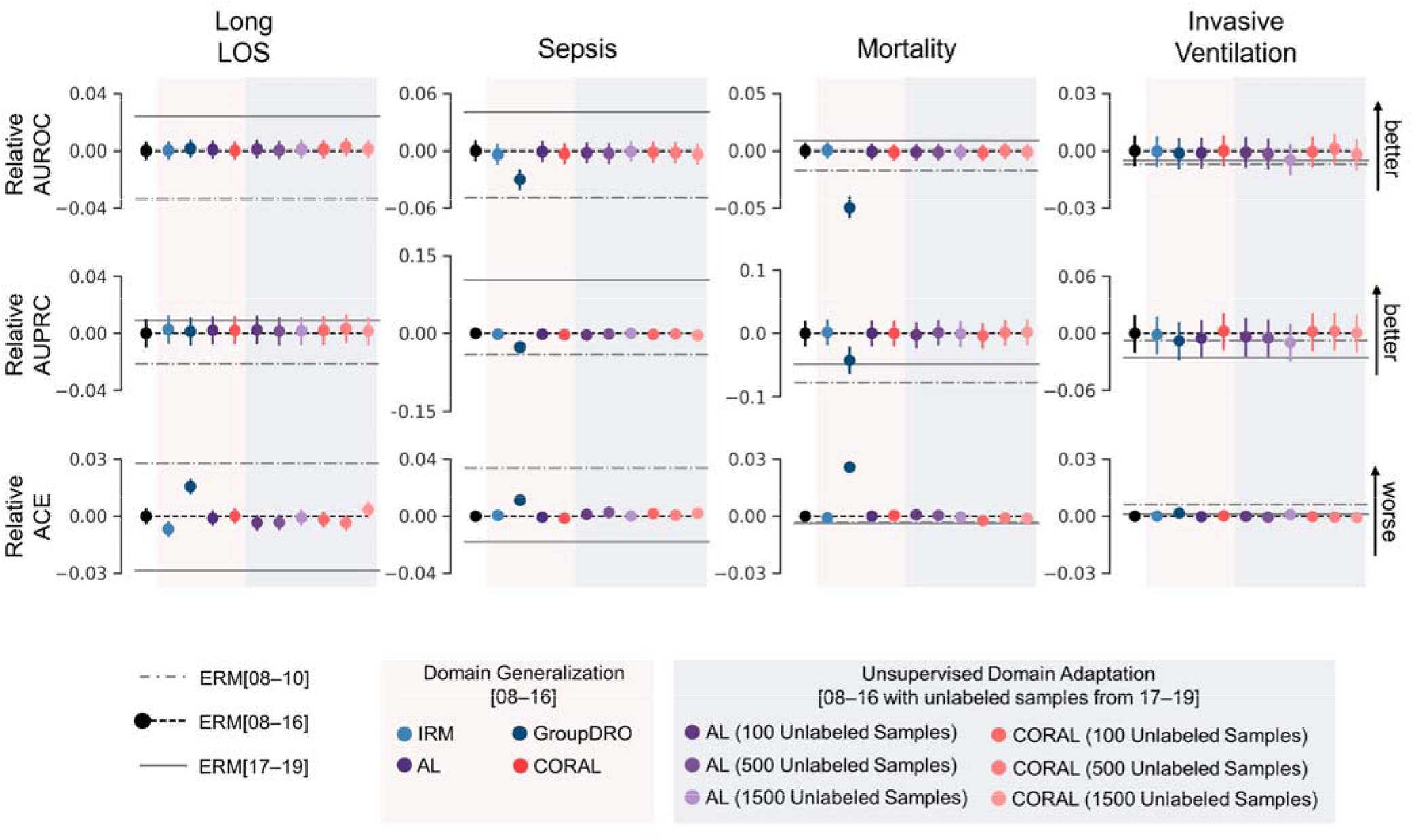
Difference in mean performance of DG and UDA approaches relative to ERM[08-16] in the target year group (2017-2019). Performance of ERM[08-10] (train set 2008-2010 and test set 2017-2019, dashed line) and ERM[17-19] (train and test sets 2017-2019, solid line) models are also shown for comparison. Error bars indicate 95% confidence interval obtained from 10,000 bootstrap iterations. Here, we show results from three of the four experimental conditions using differing number of unlabelled samples for UDA – we did not observe meaningful differences across the number of unlabelled samples evaluated. Numerical representation of the performance measures relative to ERM[08-16] are presented in **eTable 3**. Abbreviations, LOS: length of stay; ERM: empirical risk minimization; IRM: invariant risk minimization; AL: adversarial learning; GroupDRO: group distributionally robust optimization; CORAL: correlation alignment; AUROC: area under the receiver operating characteristics curve; AUPRC: area under the precision recall curve; ACE: absolute calibration error; domain generalization: DG; unsupervised domain adaptation: UDA.

## DISCUSSION

Our results revealed heterogeneity in the impact of temporal dataset shift across clinical prediction tasks, with the largest impact on sepsis prediction. When compared to ERM[08-16], DG and UDA algorithms did not substantially improve model robustness. In some cases, DG and UDA algorithms produced less performant models than ERM. We also illustrated the impact of temporal dataset shift and the effect of mitigation approaches for clinical audiences so that they can determine whether the extent of dataset shift precludes utilization in practice.

The heterogeneity of impact by temporal dataset shift as revealed by our baseline experiment echoes the mixed results in model deterioration across several studies that made predictions of clinical outcomes in various populations.^5^ This calls for careful investigation of potential model degradation due to temporal dataset shift at both the population and task level. In addition, these investigations should translate model degradation, typically measured as change in AUROC, into change in clinically relevant performance measures^27^ or utility in allocation of resources,^28^ and place its impact in the context of clinical decision making and downstream processes.^29-32^

This study is one of the first to benchmark the capability of DG and UDA algorithms on EHR data across multiple clinical prediction tasks to mitigate the impact of temporal dataset shift. Our findings align with recent empirical evaluations of DG algorithms demonstrating that they do not outperform ERM under distribution shift across data sources or hospitals in real-world clinical datasets.^33,34^ The reasons underlying the failure of DG algorithms are topics of active research, with several recent works offering theory to explain why models derived with IRM and groupDRO are typically not more robust than ERM in practice.^35,36^ Furthermore, other work has demonstrated that UDA objectives based on distribution matching, such as AL and CORAL, failed to improve generalization to the target domain under shifts in the outcome rate or in the association between the outcome and features^37,38^. These findings highlight the difficulty of improving robustness to dataset shift with purely data-driven techniques that do not leverage prior knowledge of the nature of the shift. Future research should explore methods that learn robust models by incorporating domain knowledge as to which causal mechanisms are likely to be stable or change across time^39^.

Strengths of this study include the use of multiple clinical outcomes and the illustration of temporal dataset shift and its mitigations using more clinically relevant metrics. There are several limitations in this study. First, the coarse characterization of temporal dataset shift did not offer insight about the rate at which model performance deteriorated. This was due to the deidentification in the MIMIC-IV database that left year group as the only time information that followed a correct chronological order across patients. Second, using data from the target year group to estimate best-case models is not realistic in real-world deployment as such data might not be available. Third, our assessment of clinical implications did not consider clinical use-cases in which the model alerts physicians of patients with the highest risks (i.e., acting on a threshold that is adaptively selected)^40^. In those scenarios, the amount of agreement in the ranking of risks between models need to be additionally considered.

In conclusion, DG and UDA failed to produce more robust models compared to ERM in the setting of temporal dataset shift. Alternate approaches are required to preserve model performance over time in clinical medicine.

## Data Availability

The data used for this study is publicly available. Instructions for access can be found at https://mimic.mit.edu/iv/. The code for analyses is open source and available at https://github.com/sungresearch/mimic4ds_public.

## ACKNOWEDGEMENTS

LS is the Canada Research Chair in Pediatric Oncology Supportive Care.

## AUTHOR CONTRIBUTIONS

Study concepts and design: All

Data acquisition: LLG, LS, AJ

Data analysis: LLG, SRP

Data interpretation: All

Drafting the manuscript or revising it critically for important intellectual content: All

Final approval of version to be published: All

Agreement to be accountable for all aspects of the work: All

## ETHICAL APPROVAL

As this study was conducted on de-identified electronic health records, Ethics Committee/Institutional Review Board approval was waived.

## Footnotes

a https://github.com/sungresearch/mimic4ds_public

## REFERENCES

1. Rajkomar A, Oren E, Chen K, et al. Scalable and accurate deep learning with electronic health records. NPJ Digit Med. 2018;1:18.

2. Seneviratne MG, Shah NH, Chu L. Bridging the implementation gap of machine learning in healthcare. BMJ Innovations. 2020;6:45–47.

3. Sendak MP, Balu S, Schulman KA. Barriers to Achieving Economies of Scale in Analysis of EHR Data. A Cautionary Tale. Appl Clin Inform. 2017;8(3):826–831.

4. Moreno-Torres JG, Raeder T, Alaiz-Rodríguez R, Chawla NV, Herrera F. A unifying view on dataset shift in classification. Pattern Recognit. 2012;45(1):521–530.

5. Guo LL, Pfohl SR, Fries J, et al. Systematic Review of Approaches to Preserve Machine Learning Performance in the Presence of Temporal Dataset Shift in Clinical Medicine. In:2021.

6. Davis SE, Greevy RA, Jr., Lasko TA, Walsh CG, Matheny ME. Detection of calibration drift in clinical prediction models to inform model updating. J Biomed Inform. 2020;112:103611.

7. Davis SE, Greevy RA, Fonnesbeck C, Lasko TA, Walsh CG, Matheny ME. A nonparametric updating method to correct clinical prediction model drift. J Am Med Inform Assoc. 2019;26(12):1448–1457.

8. Siregar S, Nieboer D, Versteegh MIM, Steyerberg EW, Takkenberg JJM. Methods for updating a risk prediction model for cardiac surgery: a statistical primer. Interact Cardiovasc Thorac Surg. 2019;28(3):333–338.

9. Nestor B, McDermott MBA, Boag W, et al. Feature Robustness in Non-stationary Health Records: Caveats to Deployable Model Performance in Common Clinical Machine Learning Tasks. Proceedings of the 4th Machine Learning for Healthcare Conference; 2019.

10. Zhou K, Liu Z, Qiao Y, Xiang T, Loy CC. Domain Generalization: A Survey. arXiv. 2021:1–21. https://arxiv.org/abs/2103.02503. xPublished 31 Mar 2021. Accessed 05 May 2021.

11. Wilson G, Cook DJ. A Survey of Unsupervised Deep Domain Adaptation. ArXiv. 2020. https://arxiv.org/abs/1812.02849. Accessed 21 May 21.

12. MIMIC-IV. PhysioNet; 2021. https://physionet.org/content/mimiciv/1.0/. Accessed 21 May 2021.

13. Singer M, Deutschman CS, Seymour CW, et al. The Third International Consensus Definitions for Sepsis and Septic Shock (Sepsis-3). JAMA. 2016;315(8):801–810.

14. Reps JM, Schuemie MJ, Suchard MA, Ryan PB, Rijnbeek PR. Design and implementation of a standardized framework to generate and evaluate patient-level prediction models using observational healthcare data. J Am Med Inform Assoc. 2018;25(8):969–975.

15. Varnik V. Principles of risk minimization for learning theory. Advances in Neural Information Processing Systems 4; 1991.

16. Arjovsky M, Bottou L, Gulrajani I, Lopez-Paz D. Invariant Risk Minimization. ArXiv. 2020. https://arxiv.org/abs/1907.02893. xPublished 27 Mar 2020. Accessed 21 May 21.

17. Sagawa S, Koh PW, Hashimoto TB, Liang P. Distributionally Robust Neural Networks for Group Shifts: On the Importance of Regularization for Worst-Case Generalization. ArXiv. 2020. https://arxiv.org/abs/1911.08731. xPublished 02 Apr 2020. Accessed 21 May 21.

18. Sun B, Saenko K. Deep CORAL: Correlation Alignment for Deep Domain Adaptation. European conference on computer vision; 2016; University of Massachusetts Lowell, Boston University.

19. Ganin Y, Ustinova E, Ajakan H, et al. Domain-Adversarial Training of Neural Networks. J Mach Learn Res. 2016;17:1–35.

20. Pfohl S, Marafino B, Coulet A, Rodriguez F, Pala-niappan L, Shah NH. Creating Fair Models of Atherosclerotic Cardiovascular Disease. AAAI/ACM Conference on AI, Ethics, and Society (AIES’19); 2019; Honolulu, HI, USA.

21. Pfohl SR, Foryciarz A, Shah NH. An empirical characterization of fair machine learning for clinical risk prediction. J Biomed Inform. 2021;113:103621.

22. Austin PC, Steyerberg EW. The Integrated Calibration Index (ICI) and related metrics for quantifying the calibration of logistic regression models. Stat Med. 2019;38(21):4051–4065.

23. Efron B, Tibshirani R. An introduction to the bootstrap. New York: Chapman & Hall; 1993.

24. Van Rossum G, Drake F. Python Language Reference, version 3.8. https://www.python.org/ Accessed 21 May, 2021.

25. Pedregosa F, Varoquaux G, Gramfort A, et al. Scikit-learn: Machine Learning in Python. J Mach Learn Res. 2011;12:2825–2830.

26. Paszke A, Gross S, Massa F, et al. PyTorch: An Imperative Style, High-Performance Deep Learning Library. NeurIPS. 2019;32:8024–8035.

27. Shah NH, Milstein A, Bagley Ph DS. Making Machine Learning Models Clinically Useful. JAMA. 2019;322(14):1351–1352.

28. Ko M, Chen E, Agrawal A, et al. Improving Hospital Readmission Prediction using Individualized Utility Analysis. Journal of Biomedical Informatics. 2021:103826.

29. Char DS, Shah NH, Magnus D. Implementing Machine Learning in Health Care - Addressing Ethical Challenges. N Engl J Med. 2018;378(11):981–983.

30. Morse KE, Bagley SC, Shah NH. Estimate the hidden deployment cost of predictive models to improve patient care. Nature Medicine. 2020;26(1):18–19.

31. Liu VX, Bates DW, Wiens J, Shah NH. The number needed to benefit: estimating the value of predictive analytics in healthcare. (1527-974X (Electronic)).

32. Li RC, Asch SM, Shah NH. Developing a delivery science for artificial intelligence in healthcare. npj Digital Medicine. 2020;3(1):107.

33. Koh PW, Sagawa S, Marklund H, et al. WILDS: A Benchmark of in-the-Wild Distribution Shifts. ArXiv. 2021:1–87. https://arxiv.org/abs/2012.07421.

34. Zhang H, Dullerud N, Seyyed-Kalantari L, Morris Q, Joshi S, Ghassemi M. An empirical framework for domain generalization in clinical settings. ACM Conference on Health, Inference, and Learning (ACM CHIL’21); 2021; Virtual Event.

35. Rosenfeld E, Ravikumar P, Risteski A. The Risks of Invariant Risk Minimization. ArXiv. 2020. https://arxiv.org/abs/2010.05761.

36. Rosenfeld E, Ravikumar P, Risteski A. An Online Learning Approach to Interpolation and Extrapolation in Domain Generalization. arXiv. 2021.

37. Wu Y, Winston E, Kaushik D, Lipton Z. Domain Adaptation with Asymmetrically-Relaxed Distribution Alignment. Proceedings of the 36th International Conference on Machine Learning; 2019; Proceedings of Machine Learning Research.

38. Zhao H, Combes RTD, Zhang K, Gordon G. On Learning Invariant Representations for Domain Adaptation. 36th International Conference on Machine Learning, ICML 2019; 2019.

39. Subbaswamy A, Schulam P, Saria S. Preventing Failures Due to Dataset Shift: Learning Predictive Models That Transport. Proceedings of the Twenty-Second International Conference on Artificial Intelligence and Statistics; 2019; Proceedings of Machine Learning Research.

40. Manz CR, Parikh RB, Small DS, et al. Effect of Integrating Machine Learning Mortality Estimates With Behavioral Nudges to Clinicians on Serious Illness Conversations Among Patients With Cancer: A Stepped-Wedge Cluster Randomized Clinical Trial. JAMA Oncol. 2020;6(12):e204759.

