## Supplemental Document for "Evaluation of Domain Generalization and Adaptation on Improving Model Robustness to Temporal Dataset Shift in Clinical Medicine"

**Supplementary Content**

| Pages |  |  |
| --- | --- | --- |
| 2-10 | **eMethods** | Additional details on clinical outcomes, feature extraction, models and learning algorithms |
| 11 | **eReferences** |  |
| 12 | **eFigure 1** | AUROC in the test set of 2017-2019 across hyperparameter values of DG and UDA learning algorithms |
| 13 | **eFigure 2** | AUPRC in the test set of 2017-2019 across hyperparameter values of DG and UDA learning algorithms |
| 14 | **eFigure 3** | ACE in the test set of 2017-2019 across hyperparameter values of DG and UDA learning algorithms |
| 15 | **eTable 1** | Difference in the performance measures of DG, UDA, and ERM[08-16] models relative to ERM[08-10] 2017–2019 |
| 16 | **eTable 2** | Difference in the performance measures of DG, UDA, and ERM[08-16] models relative to ERM[17-19] in 2017–2019 |
| 17 | **eTable 3** | Difference in the performance measures of DG and UDA models relative to ERM[08-16] in 2017-2019 |
| 18 | **eTable 4** | Performance measures of ERM[08-16], DG, and UDA models in the test sets of 2008–2016 |
| 19 | **eTable 5** | Threshold-based metrics in the sepsis prediction task across different thresholds |

**eMethods.** Additional details on clinical outcomes, feature extraction, models and learning algorithms.

I. Clinical outcomes
II. Number of clinical features extracted from MIMIC-IV for each category and time interval
III. Model hyperparameters
IV. Learning algorithm objective and hyperparameter
V. Hyperparameter selection and model training
VI. Selected model hyperparameters
VII. Selected algorithm hyperparameter

**I. Clinical outcomes**

| **Clinical outcome** | **Description** |
| --- | --- |
| Mortality | Positive label was defined as in-hospital mortality within 7 days from the time of prediction. |
| Long LOS | Positive label was defined as ICU stay for more than 3 days from the time of prediction. |
| Invasive  Ventilation | Positive label was defined as initiation of invasive ventilation within 24 hours from the time of prediction. Invasive ventilation was defined as receiving mechanical ventilation through a tracheostomy tube, an endotracheal tube, or using a list of ventilator modes included in the concept definition of invasive ventilation in the MIMIC-IV GitHub repository^a^. |
| Sepsis | Positive label was defined as developing sepsis within 7 days from the time of prediction. Sepsis was defined according to the Sepsis-3 criteria^1^ implemented in the MIMIC-IV GitHub repository. Specifically, the patient met sepsis criteria if the patient was suspected to have an infection and had an increase in SOFA score of 2 or more relative to the baseline, which was assumed to be 0 prior to ICU admission. Suspected infection was defined as concomitant administration of systemic antibiotics and sampling of cultures. Events were considered concomitant if antibiotics were administered within 72 hours from the sampling of cultures, or if cultures were sampled within 24 hours from the administration of antibiotics. The time of suspected infection was defined as the time of antibiotics administration or culture sampling, whichever occurred first. The onset time of sepsis was defined as the time when the patient was suspected of infection and had a SOFA score of 2 or more. |

*All clinical outcomes were binary. The time of prediction was set as 4 hours after the time of ICU admission. Patients that received invasive ventilation or developed sepsis according to our definition prior to the time of prediction were excluded.

Abbreviations. LOS: length of stay; SOFA: Sequential Organ Failure Assessment; ICU: intensive care unit.

^a^ https://github.com/MIT-LCP/mimic-iv

**II. Number of clinical features extracted from MIMIC-IV for each category and time interval**

|  |  | Count  Features^a^ | | | | |  | Measurement Features^b^ | |
| --- | --- | --- | --- | --- | --- | --- | --- | --- | --- |
|  |  | Diagnosis  (ICD) | Procedure  (ICD) | Procedure  (HCPCS) | Prescription  (NDC) | Labs  (item ID) |  | Lab test results | ICU charts |
| Mortality / Long Length of Stay (total features extracted = 48423)^c^ | | | | | | | | | |
|  | all to -180 d | 2189 | 545 | 42 | 2037 | 556 |  | 1071 | 0 |
|  | -180 d to -30 d | 5450 | 1727 | 198 | 3424 | 729 |  | 1395 | 0 |
|  | -30 d to -7 d | 4534 | 1402 | 121 | 3390 | 714 |  | 1326 | 0 |
|  | -7 d to 0 | 3262 | 900 | 68 | 3657 | 743 |  | 1389 | 0 |
|  | 0 to 4 h | 0 | 0 | 0 | 3154 | 653 |  | 1257 | 2490 |
| Invasive Ventilation (total features extracted = 44907) | | | | | | | | | |
|  | all to -180 d | 1943 | 467 | 34 | 1885 | 537 |  | 1026 | 0 |
|  | -180 d to -30 d | 4922 | 1524 | 176 | 3262 | 717 |  | 1365 | 0 |
|  | -30 d to -7 d | 4091 | 1190 | 106 | 3221 | 698 |  | 1287 | 0 |
|  | -7 d to 0 | 2909 | 765 | 58 | 3493 | 731 |  | 1374 | 0 |
|  | 0 to 4 h | 0 | 0 | 0 | 2985 | 634 |  | 1212 | 2295 |
| Sepsis (total features extracted = 40691) | | | | | | | | | |
|  | all to -180 d | 1581 | 362 | 31 | 1635 | 482 |  | 930 | 0 |
|  | -180 d to -30 d | 4269 | 1310 | 151 | 3031 | 690 |  | 1323 | 0 |
|  | -30 d to -7 d | 3459 | 961 | 78 | 3007 | 688 |  | 1287 | 0 |
|  | -7 d to 0 | 2473 | 614 | 42 | 3413 | 718 |  | 1335 | 0 |
|  | 0 to 4 h | 0 | 0 | 0 | 2792 | 612 |  | 1146 | 2271 |

*All time intervals were relative to the time of ICU admission. Diagnosis and procedure codes were not obtained 0 to 4 hours after admission time because ICD and HCPCS codes are assigned at discharge time.

Abbreviations. ICD: International Classification of Disease; HCPCS: Healthcare Common Procedure Coding System; NDC: National Drug Codes; ICU: intensive care unit.

^a^Counts of unique concept identifiers.

^b^Summary statistics (min, mean, max) of numerical values from lab test results and ICU charts in each time interval.

^c^Mortality and long length of stay have the same number of features extracted because they share the same cohort. In comparison, invasive ventilation and sepsis have smaller cohort sizes due to the exclusion of patients with those outcomes prior to prediction time (4 hours after ICU admission).

**III. Model hyperparameters**

| Hyperparameter | | Values |
| --- | --- | --- |
| To be selected | |  |
|  | Dropout probability | 0, 0.25, 0.5, 0.75 |
|  | Gamma^a^ | 1, 0.95 |
|  | Hidden dimension^b^ | 128, 256 |
|  | Learning rate | 0.01, 0.0001, 0.000001 |
|  | Number of hidden layers | 2, 4, 6 |
| Fixed^c^ | |  |
|  | Optimizer | Adam |
|  | Training iterations | 200 |
|  | Early stopping | True |
|  | Early stopping patience | 25 |
|  | Batch size^c^ | 256 |
|  | Number of batches per epoch | 100 |

^a^Gamma is the multiplicative factor that determines the amount of exponential decay in learning rate every epoch.

^b^Our fully connected feedforward neural network models have fixed widths determined by *hidden dimension* and the same dropout probability for each hidden layer determined by *dropout probability* across n layers, where n is determined by the *number of hidden layers*.

**IV. Learning algorithm objective and hyperparameter**

| Learning Algorithm | Objective and Hyperparameter | Hyperparameter values |
| --- | --- | --- |
| Empirical Risk Minimization^2^  (ERM) | ERM seeks to minimize average training loss (in our case the cross-entropy) across all training environments. There are no algorithm-specific hyperparameters for ERM. | - |
| Invariant Risk Minimization^3^  (IRM) | IRM assumes the existence of a latent representation (i.e., hidden layer activations) on which the optimal classifier is the same for every training environment. To approximate this representation, IRM regularizes the loss (in our case the cross-entropy loss) by the squared norm of the gradients of the loss with respect to the parameters of a fixed classifier on top of the representation for each training environment. The hyperparameter for the regularization, λ, is the multiplicative factor that modulates the impact of IRM penalty on the overall learning objective (i.e., minimize average cross entropy + λ * IRM penalty). | 10^-1^, 10^-0^, 10^1^, 10^2^, 10^3^, 10^4^ |
| Group Distributionally Robust Optimization^4^ (GroupDRO) | GroupDRO seeks to improve worst-case performance by performing ERM while increasing the importance of the training environments with larger average training loss (e.g., cross-entropy loss). As such, GroupDRO does not “learn” any invariance. The hyperparameter η is the learning rate of the exponentiated gradient descent update on the weights over the environment losses. | 10^-1^, 10^-2^, 10^-3^, 10^-4^, 10^-5^, 10^-6^ |
| Domain Adversarial Learning^5^  (AL) | AL matches the distribution of the data encoded in the latent space across environments using an adversarial network and regularizes the loss (in our case the cross-entropy loss) with an adversarial objective that minimizes discriminability between environments. The hyperparameter for the regularization, λ, is the multiplicative factor that modulates the impact of the adversarial objective on the overall learning objective. In our implementation of AL we opted to perform early stopping and model selection on the basis of the the average cross-entropy. | 10^0^, 10^-1^, 10^-2^, 10^-3^, 10^-4^, 10^-5^ |
| Correlation Alignment  (CORAL)^6^ | CORAL matches the mean and covariance of the distribution of the data encoded in the latent space across environments. To that end, CORAL regularizes the loss (in our case the cross-entropy loss) by the differences in the means and covariances of the distributions. The hyperparameter for the regularization, λ, is the multiplicative factor that modulates the impact of the CORAL penalty on the overall learning objective (i.e., minimize average cross entropy + λ * CORAL penalty). | 10^-2^, 10^-1^, 10^0^, 10^1^, 10^2^, 10^3^ |

*Range of hyperparameter values were selected based on previous benchmark tasks.^7,8^

Abbreviations. DG: domain generalization; UDA: unsupervised domain adaptation; IRM: invariant risk minimization; GroupDRO: group distributionally robust optimization; AL: adversarial learning; CORAL: correlational alignment.

**V. Hyperparameter Selection and Model Training**

For ERM models, we conducted 100 random grid-searches (see section III for model hyperparameters) and selected hyperparameters based on mean validation cross-entropy loss. For DG and UDA models, we set the model hyperparameters to those selected for ERM. Then, we selected algorithm hyperparameters based on the algorithm-specific objective evaluated in the validation set (see section IV for algorithm objective and hyperparameter). Sections VI and VII list the selected hyperparameters for models and algorithms, respectively.

Using the selected hyperparameters, we trained 20 NN models with different initializations on the training sets of each experiment for each combination of task and experiment-specific characteristic (e.g., year group in the baseline experiment). Model training was performed using the Adam optimizer^9^ for up to 200 iterations of 100 batches (256 samples per batch) and stopped early if no improvement in the algorithm-specific objective evaluated in the validation set was observed for 25 iterations.

Hyperparameter sweep for DG and UDA algorithms:

We recognized that our hyperparameter selection procedure for DG and UDA algorithms might encourage the selection of small λ values such that the loss function is near equivalent to ERM, and result in producing ERM-like models. Therefore, we trained additional models using the full range of algorithm hyperparameters and evaluated these models in the test set of 2017-2019 to investigate if the benefits of DG and UDA algorithms increase at higher hyperparameter values. AUROC, AUPRC, and ACE of these additional models are presented in eFigures1, 2, and 3, respectively.

*Abbreviations: ERM: empirical risk minimization; AL: domain adversarial learning; DG: domain generalization; UDA: unsupervised domain adaptation; NN: fully connected feedforward neural network models. AUROC: area under the receiver operational characteristic curve; AUPRC: area under the precision recall curve; ACE: absolute calibration error.

VI. Selected model hyperparameters

| Experiment | Clinical  Prediction  Task | Year-Group | No. Hidden Layers | Hidden Layer Dimension | Dropout Probability | Learning Rate (LR) | Exponential LR Decay Factor (Gamma) |
| --- | --- | --- | --- | --- | --- | --- | --- |
| Baseline | Mortality | 2008 - 2010 | 2 | 256 | 0.25 | 1e-05 | 0.95 |
| Baseline | Mortality | 2011 - 2013 | 2 | 256 | 0.5 | 1e-04 | 0.95 |
| Baseline | Mortality | 2014 - 2016 | 2 | 128 | 0.5 | 1e-05 | 1.0 |
| Baseline | Mortality | 2017 - 2019 | 2 | 256 | 0.25 | 1e-05 | 0.95 |
| Baseline | Long LOS | 2008 - 2010 | 2 | 128 | 0.0 | 1e-05 | 0.95 |
| Baseline | Long LOS | 2011 - 2013 | 6 | 256 | 0.25 | 1e-05 | 0.95 |
| Baseline | Long LOS | 2014 - 2016 | 2 | 128 | 0.5 | 1e-05 | 1.0 |
| Baseline | Long LOS | 2017 - 2019 | 4 | 128 | 0.25 | 0.0001 | 0.95 |
| Baseline | Invasive Ventilation | 2008 - 2010 | 2 | 256 | 0.0 | 1e-05 | 0.95 |
| Baseline | Invasive Ventilation | 2011 - 2013 | 2 | 256 | 0.0 | 1e-05 | 1.0 |
| Baseline | Invasive Ventilation | 2014 - 2016 | 6 | 128 | 0.25 | 1e-05 | 1.0 |
| Baseline | Invasive Ventilation | 2017 - 2019 | 2 | 128 | 0.25 | 1e-05 | 0.95 |
| Baseline | Sepsis | 2008 - 2010 | 2 | 128 | 0.25 | 1e-05 | 1.0 |
| Baseline | Sepsis | 2011 - 2013 | 4 | 128 | 0.25 | 1e-05 | 1.0 |
| Baseline | Sepsis | 2014 - 2016 | 6 | 128 | 0.0 | 1e-05 | 0.95 |
| Baseline | Sepsis | 2017 - 2019 | 6 | 128 | 0.0 | 1e-05 | 1.0 |
| DG / UDA | Mortality | 2008 - 2016 | 2 | 256 | 0.5 | 1e-05 | 1.0 |
| DG / UDA | Long LOS | 2008 - 2016 | 2 | 128 | 0.5 | 1e-05 | 1.0 |
| DG / UDA | Invasive Ventilation | 2008 - 2016 | 2 | 128 | 0.25 | 1e-05 | 0.95 |
| DG / UDA | Sepsis | 2008 - 2016 | 6 | 128 | 0.25 | 1e-05 | 1.0 |

*Abbreviations: LOS: length of stay; DG: domain generalization; UDA: unsupervised domain adaptation

VII. Selected algorithm hyperparameters

| Prediction  Task | Framework (Setting) | Method | No. Samples from Target Year Group | Hyperparameter Value |
| --- | --- | --- | --- | --- |
| Mortality | DG | AL | - | 1e-05 |
| Mortality | DG | CORAL | - | 0.1 |
| Mortality | DG | IRM | - | 0.1 |
| Mortality | DG | GroupDRO | - | 0.01 |
| Long LOS | DG | AL | - | 0.0001 |
| Long LOS | DG | CORAL | - | 10 |
| Long LOS | DG | IRM | - | 1 |
| Long LOS | DG | GroupDRO | - | 0.1 |
| Invasive Ventilation | DG | AL | - | 0.0001 |
| Invasive Ventilation | DG | CORAL | - | 10 |
| Invasive Ventilation | DG | IRM | - | 0.1 |
| Invasive Ventilation | DG | GroupDRO | - | 0.01 |
| Sepsis | DG | AL | - | 0.0001 |
| Sepsis | DG | CORAL | - | 1 |
| Sepsis | DG | IRM | - | 0.1 |
| Sepsis | DG | GroupDRO | - | 0.1 |
| Mortality | UDA | AL | 100 | 0.0001 |
| Mortality | UDA | AL | 500 | 1e-05 |
| Mortality | UDA | AL | 1000 | 0.0001 |
| Mortality | UDA | AL | 1500 | 1e-05 |
| Mortality | UDA | CORAL | 100 | 1 |
| Mortality | UDA | CORAL | 500 | 0.1 |
| Mortality | UDA | CORAL | 1000 | 0.1 |
| Mortality | UDA | CORAL | 1500 | 0.1 |
| Long LOS | UDA | AL | 100 | 0.001 |
| Long LOS | UDA | AL | 500 | 1e-05 |
| Long LOS | UDA | AL | 1000 | 0.01 |
| Long LOS | UDA | AL | 1500 | 1e-05 |
| Long LOS | UDA | CORAL | 100 | 0.01 |
| Long LOS | UDA | CORAL | 500 | 0.1 |
| Long LOS | UDA | CORAL | 1000 | 0.01 |
| Long LOS | UDA | CORAL | 1500 | 10 |
| Invasive Ventilation | UDA | AL | 100 | 0.001 |
| Invasive Ventilation | UDA | AL | 500 | 0.0001 |
| Invasive Ventilation | UDA | AL | 1000 | 0.001 |
| Invasive Ventilation | UDA | AL | 1500 | 0.01 |
| Invasive Ventilation | UDA | CORAL | 100 | 1 |
| Invasive Ventilation | UDA | CORAL | 500 | 0.1 |
| Invasive Ventilation | UDA | CORAL | 1000 | 10 |
| Invasive Ventilation | UDA | CORAL | 1500 | 1 |
| Sepsis | UDA | AL | 100 | 1e-05 |
| Sepsis | UDA | AL | 500 | 0.01 |
| Sepsis | UDA | AL | 1000 | 1e-05 |
| Sepsis | UDA | AL | 1500 | 1e-05 |
| Sepsis | UDA | CORAL | 100 | 0.01 |
| Sepsis | UDA | CORAL | 500 | 0.1 |
| Sepsis | UDA | CORAL | 1000 | 0.01 |
| Sepsis | UDA | CORAL | 1500 | 0.01 |

*Abbreviations: DG: domain generalization; UDA: unsupervised domain adaptation; LOS: length of stay; IRM: invariant risk minimization; GroupDRO: group distributionally robust optimization; AL: adversarial learning; CORAL: correlational alignment.

**eReferences.**

1.	Singer M, Deutschman CS, Seymour CW, et al. The Third International Consensus Definitions for Sepsis and Septic Shock (Sepsis-3). *JAMA.* 2016;315(8):801-810.

2.	Varnik V. Principles of risk minimization for learning theory. Advances in Neural Information Processing Systems 4; 1991.

3.	Arjovsky M, Bottou L, Gulrajani I, Lopez-Paz D. Invariant Risk Minimization. *ArXiv.* 2020. <https://arxiv.org/abs/1907.02893>. Published 27 Mar 2020. Accessed 21 May 21.

4.	Sagawa S, Koh PW, Hashimoto TB, Liang P. Distributionally Robust Neural Networks for Group Shifts: On the Importance of Regularization for Worst-Case Generalization. *ArXiv.* 2020. <https://arxiv.org/abs/1911.08731>. Published 02 Apr 2020. Accessed 21 May 21.

5.	Ganin Y, Ustinova E, Ajakan H, et al. Domain-Adversarial Training of Neural Networks. *J Mach Learn Res.* 2016;17:1-35.

6.	Sun B, Saenko K. Deep CORAL: Correlation Alignment for Deep Domain Adaptation. European conference on computer vision; 2016; University of Massachusetts Lowell, Boston University.

7.	Gulrajani I, Lopez-Paz D. In Search of Lost Domain Generalization. *ArXiv.* 2020. <https://arxiv.org/abs/2007.01434>.

8.	Koh PW, Sagawa S, Marklund H, et al. WILDS: A Benchmark of in-the-Wild Distribution Shifts. *ArXiv.* 2021:1-87. <https://arxiv.org/abs/2012.07421>.

9.	Kingma DP, Ba J. Adam: A Method for Stochastic Optimization. *ArXiv.* 2017. <https://arxiv.org/abs/1412.6980>.

**eFigure 1.** AUROC in the test set of 2017-2019 across hyperparameter values (section IV in eMethods) for DG and UDA learning algorithms. We observed that domain generalization and adaptation algorithms performed similar to ERM models. Furthermore, better performance were generally observed with smaller hyperparameter values. ERM[17-19] reflects ERM models trained using 2017-2019. ERM[08-16] reflects ERM models trained using 2008-2016. Abbreviations: AUROC: area under the receiver operator characteristic curve; LOS: length of stay; ERM: empirical risk minimization; IRM: invariant risk minimization; GroupDRO: group distributionally robust optimization; AL: adversarial learning; CORAL: correlational alignment.
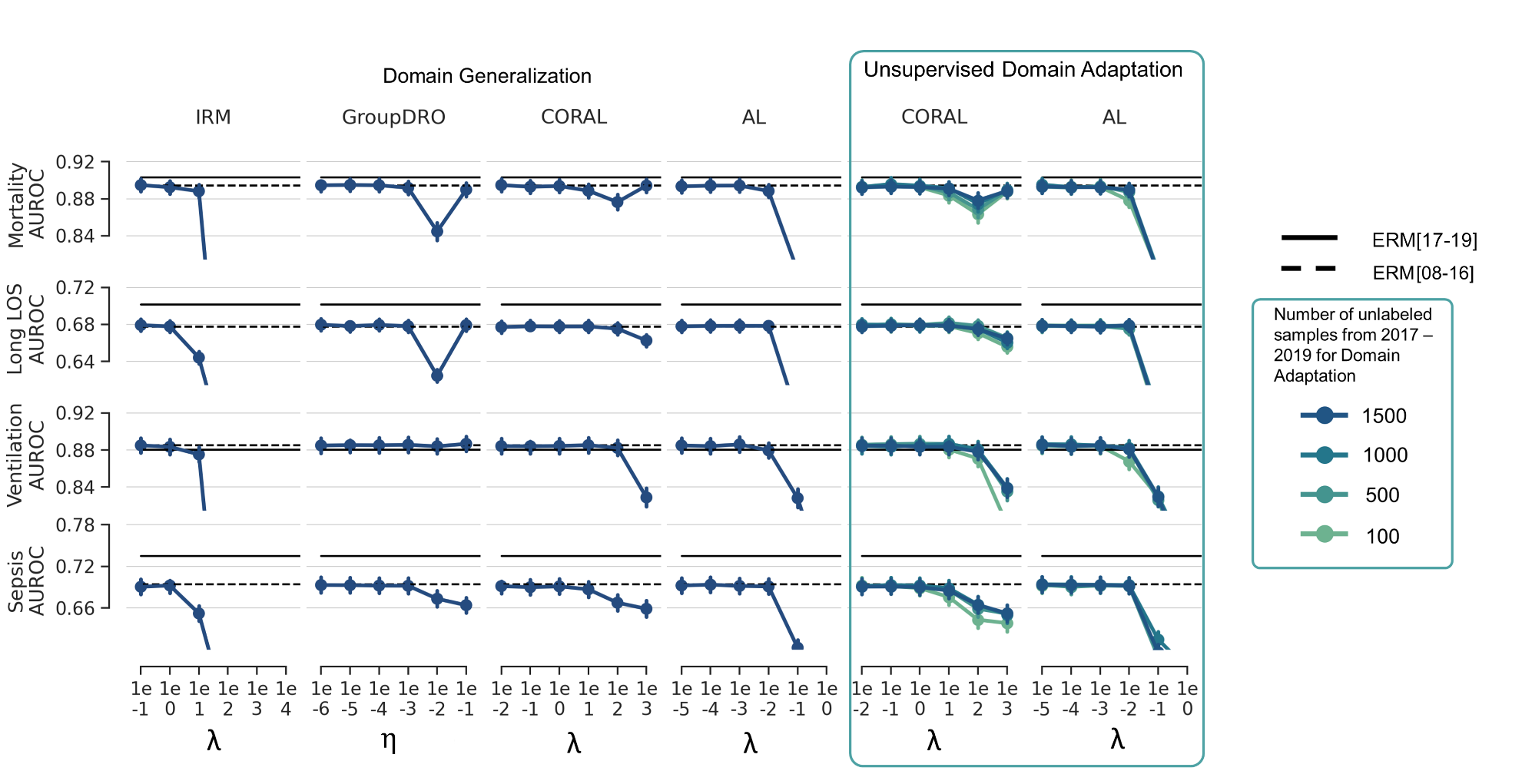

**eFigure 2.** AUPRC in the test set of 2017-2019 across hyperparameter values (section IV in eMethods) for DG and UDA learning algorithms. We observed that domain generalization and adaptation algorithms performed similar to ERM models. Furthermore, better performance were generally observed with smaller hyperparameter values. ERM[17-19] reflects ERM models trained using 2017-2019. ERM[08-16] reflects ERM models trained using 2008-2016. Abbreviations: AUPRC: area under the precision recall curve; LOS: length of stay; ERM: empirical risk minimization; IRM: invariant risk minimization; GroupDRO: group distributionally robust optimization; AL: adversarial learning; CORAL: correlational alignment.
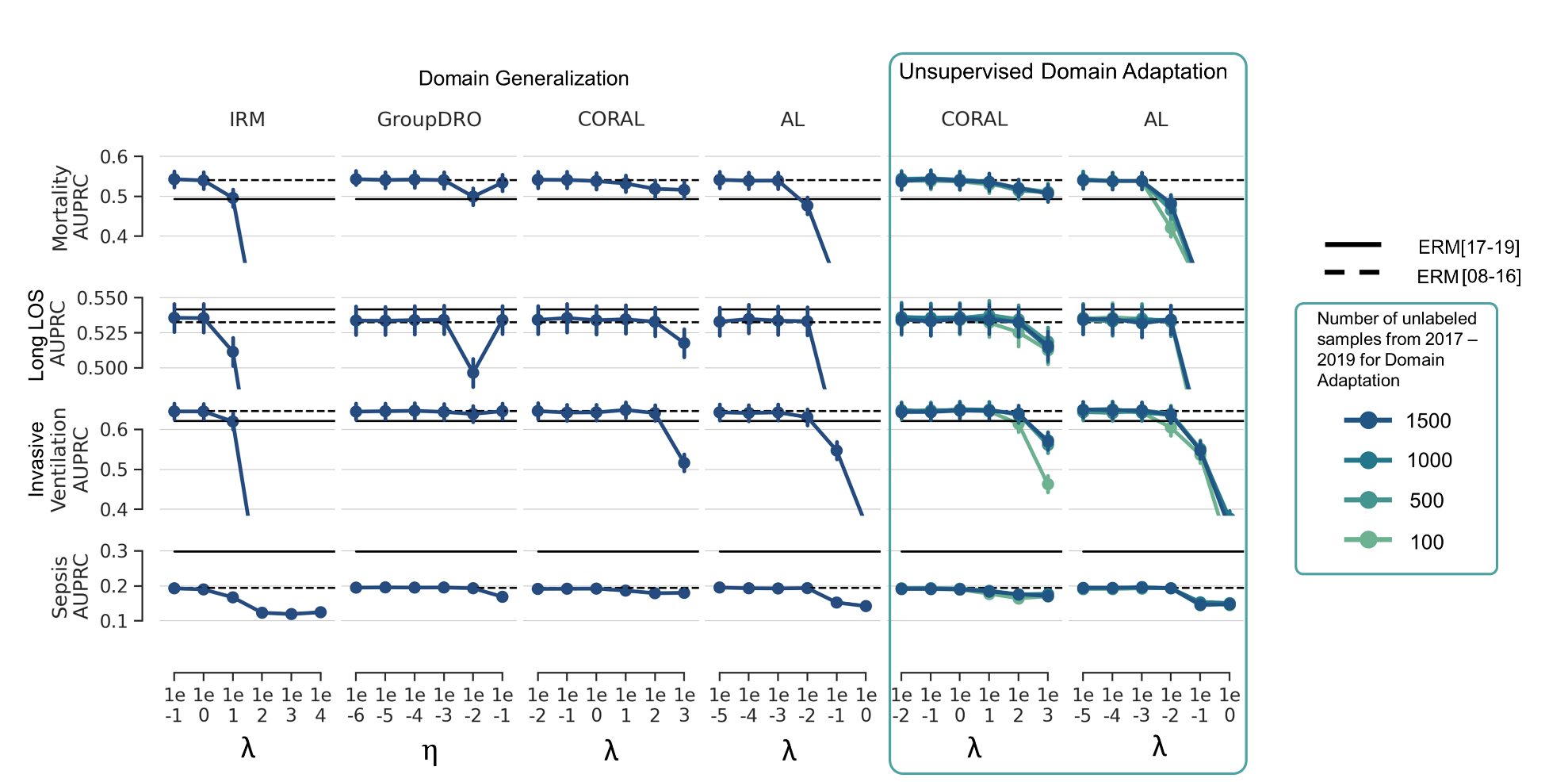

**eFigure 3.** Calibration performance (absolute calibration error) in the test set of 2017-2019 across hyperparameter values (section IV in eMethods) for DG and UDA learning algorithms. We observed that domain generalization and adaptation algorithms generally performed at similar levels as ERM[08-16] models, with one exception in which IRM led to a relatively large improvement in calibration with λ=10 in the Long LOS prediction task. ERM[17-19] reflects ERM models trained using 2017-2019. ERM[08-16] reflects ERM models trained using 2008-2016. Abbreviations: LOS: length of stay; ERM: empirical risk minimization; IRM: invariant risk minimization; GroupDRO: group distributionally robust optimization; AL: adversarial learning; CORAL: correlational alignment.

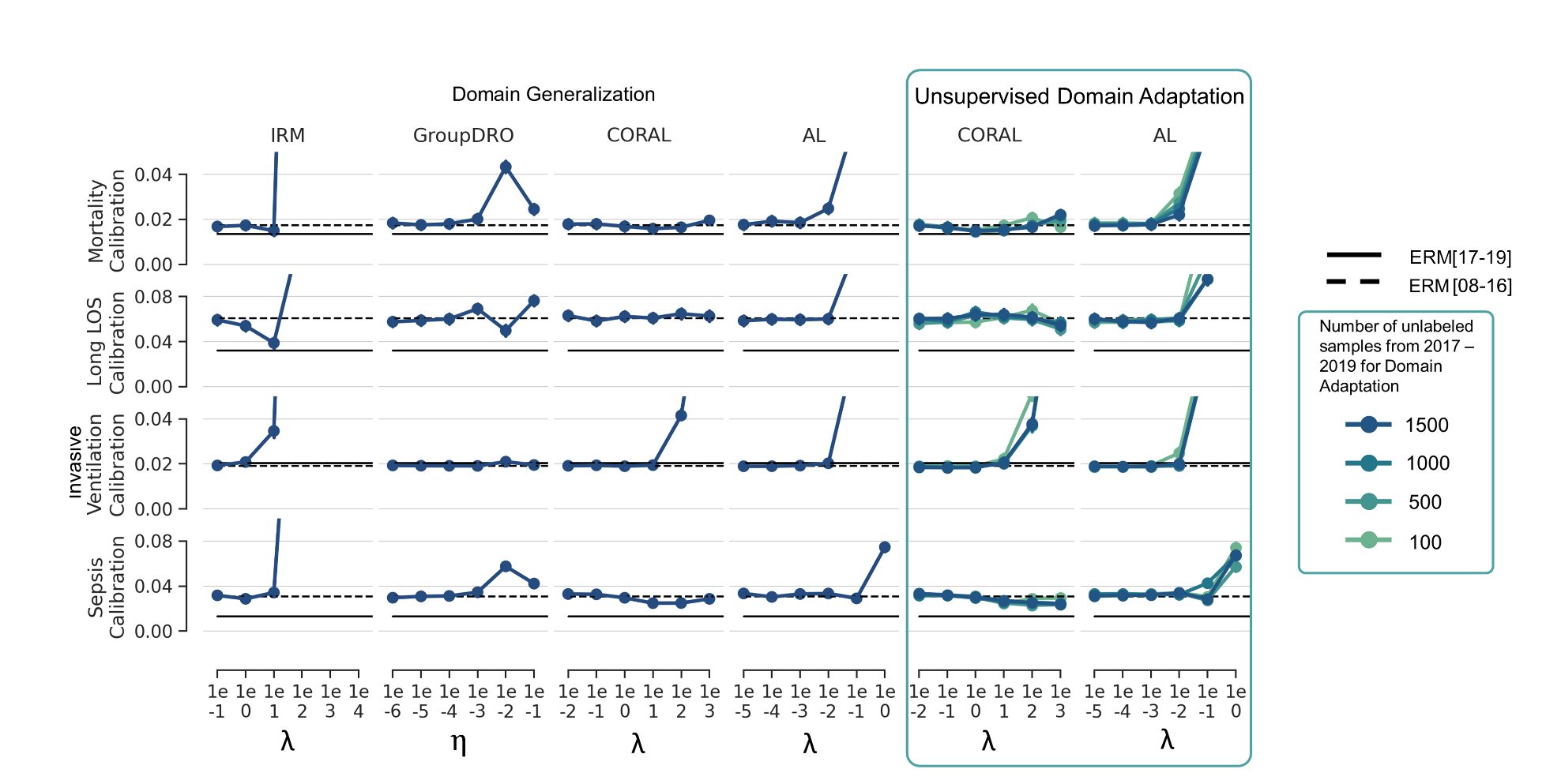

**eTable 1.** Difference in the performance measures of DG, UDA, and ERM[08-16] models relative to ERM[08-10] in 2017–2019.

|  |  | **Med (95%CI)** | **Med Difference (95% CI)** | | | | | | | | | | | | | | |
| --- | --- | --- | --- | --- | --- | --- | --- | --- | --- | --- | --- | --- | --- | --- | --- | --- | --- |
|  |  |  |  | | **Domain Generalization** | | | | | **Unsupervised Domain Adaptation** | | | | | | | |
|  |  |  |  |  | |  |  | |  | **100 OOD Samples** | | **500 OOD Samples** | | **1000 OOD Samples** | | **1500 OOD Samples** | |
|  | **Metric** | **ERM**  **[08–10]** | **ERM**  **[08-16]** | **IRM** | | **GroupDRO** | | **AL** | **CORAL** | **AL** | **CORAL** | **AL** | **CORAL** | **AL** | **CORAL** | **AL** | **CORAL** |
| **Long LOS** | | |  |  | |  |  | |  |  |  |  |  |  |  |  |  |
|  | **AUROC** | ***0.644 (0.637,0.650)*** | 0.034* (0.030,0.038) | 0.034* (0.030,0.038) | | 0.035* (0.031,0.039) | | 0.035* (0.031,0.038) | 0.034* (0.030,0.038) | 0.035* (0.031,0.039) | 0.035* (0.031,0.039) | 0.034* (0.031,0.038) | 0.036* (0.033,0.040) | 0.035* (0.031,0.039) | 0.036* (0.032,0.040) | 0.035* (0.031,0.039) | 0.035* (0.031,0.039) |
|  | **AUPRC** | ***0.512 (0.501,0.522)*** | 0.021* (0.016,0.026) | 0.024* (0.019,0.029) | | 0.022* (0.017,0.028) | | 0.023* (0.018,0.028) | 0.023* (0.018,0.028) | 0.023* (0.018,0.029) | 0.023* (0.018,0.028) | 0.023* (0.017,0.028) | 0.024* (0.019,0.030) | 0.023* (0.018,0.028) | 0.025* (0.020,0.030) | 0.023* (0.018,0.028) | 0.023* (0.017,0.028) |
|  | **ACE** | ***0.089 (0.084,0.093)*** | -0.028* (-0.030,-0.026) | -0.035* (-0.037,-0.033) | | -0.012* (-0.015,-0.010) | | -0.029* (-0.031,-0.027) | -0.028* (-0.030,-0.026) | -0.031* (-0.033,-0.029) | -0.030* (-0.032,-0.028) | -0.031* (-0.033,-0.029) | -0.031* (-0.033,-0.029) | -0.030* (-0.032,-0.028) | -0.028* (-0.030,-0.026) | -0.029* (-0.031,-0.026) | -0.024* (-0.026,-0.022) |
| **Sepsis** | | |  |  | |  |  | |  |  |  |  |  |  |  |  |  |
|  | **AUROC** | ***0.645 (0.634,0.656)*** | 0.049* (0.041,0.057) | 0.045* (0.038,0.053) | | 0.019* (0.013,0.024) | | 0.048* (0.041,0.056) | 0.046* (0.038,0.054) | 0.047* (0.039,0.055) | 0.048* (0.040,0.055) | 0.046* (0.038,0.054) | 0.047* (0.039,0.055) | 0.049* (0.041,0.057) | 0.046* (0.039,0.054) | 0.048* (0.040,0.056) | 0.046* (0.038,0.053) |
|  | **AUPRC** | ***0.155 (0.148,0.164)*** | 0.039* (0.034,0.045) | 0.037* (0.032,0.043) | | 0.013* (0.009,0.017) | | 0.038* (0.032,0.043) | 0.037* (0.031,0.042) | 0.036* (0.031,0.042) | 0.037* (0.032,0.043) | 0.038* (0.032,0.044) | 0.038* (0.033,0.044) | 0.038* (0.033,0.044) | 0.037* (0.032,0.043) | 0.039* (0.034,0.046) | 0.035* (0.030,0.041) |
|  | **ACE** | ***0.065 (0.061,0.068)*** | -0.034* (-0.036,-0.031) | -0.033* (-0.036,-0.031) | | -0.023* (-0.025,-0.021) | | -0.035* (-0.037,-0.032) | -0.035* (-0.038,-0.033) | -0.032* (-0.035,-0.030) | -0.032* (-0.035,-0.029) | -0.031* (-0.034,-0.028) | -0.033* (-0.036,-0.030) | -0.032* (-0.035,-0.029) | -0.033* (-0.036,-0.031) | -0.034* (-0.036,-0.031) | -0.032* (-0.034,-0.029) |
| **Mortality** | | |  |  | |  |  | |  |  |  |  |  |  |  |  |  |
|  | **AUROC** | ***0.877 (0.869,0.885)*** | 0.017* (0.013,0.021) | 0.017* (0.013,0.022) | | -0.033* (-0.040,-0.026) | | 0.016* (0.012,0.020) | 0.016* (0.011,0.020) | 0.016* (0.011,0.020) | 0.015* (0.011,0.019) | 0.016* (0.012,0.020) | 0.017* (0.013,0.021) | 0.015* (0.011,0.019) | 0.019* (0.015,0.023) | 0.016* (0.012,0.020) | 0.016* (0.012,0.020) |
|  | **AUPRC** | ***0.464 (0.443,0.486)*** | 0.077* (0.066,0.089) | 0.079* (0.067,0.090) | | 0.035* (0.019,0.050) | | 0.077* (0.065,0.089) | 0.077* (0.065,0.089) | 0.074* (0.062,0.086) | 0.073* (0.061,0.085) | 0.078* (0.066,0.090) | 0.078* (0.066,0.090) | 0.074* (0.062,0.086) | 0.081* (0.069,0.092) | 0.076* (0.064,0.088) | 0.079* (0.067,0.091) |
|  | **ACE** | ***0.014 (0.013,0.016)*** | 0.003* (0.002,0.004) | 0.002* (0.001,0.003) | | 0.029* (0.027,0.031) | | 0.003* (0.002,0.004) | 0.004* (0.002,0.005) | 0.004* (0.003,0.005) | 0.001 (-0.001,0.002) | 0.004* (0.002,0.005) | 0.002* (0.001,0.003) | 0.003* (0.002,0.004) | 0.002* (0.000,0.003) | 0.003* (0.001,0.004) | 0.002* (0.001,0.003) |
| **Invasive Ventilation** | | |  |  | |  |  | |  |  |  |  |  |  |  |  |  |
|  | **AUROC** | ***0.878 (0.870,0.886)*** | 0.007* (0.002,0.012) | 0.007* (0.002,0.011) | | 0.006* (0.001,0.011) | | 0.006* (0.001,0.011) | 0.007* (0.002,0.012) | 0.006* (0.001,0.011) | 0.006* (0.002,0.011) | 0.005* (0.001,0.010) | 0.008* (0.003,0.013) | 0.007* (0.002,0.011) | 0.008* (0.004,0.013) | 0.003 (-0.002,0.007) | 0.005* (0.000,0.010) |
|  | **AUPRC** | ***0.639 (0.619,0.659)*** | 0.007 (-0.004,0.018) | 0.006 (-0.005,0.017) | | -0.001 (-0.012,0.011) | | 0.002 (-0.009,0.013) | 0.009 (-0.002,0.021) | 0.003 (-0.008,0.015) | 0.009 (-0.002,0.020) | 0.002 (-0.010,0.014) | 0.009 (-0.002,0.021) | 0.003 (-0.008,0.014) | 0.010 (-0.001,0.021) | -0.002 (-0.014,0.009) | 0.008 (-0.004,0.019) |
|  | **ACE** | ***0.025 (0.023,0.027)*** | -0.006* (-0.008,-0.004) | -0.006* (-0.008,-0.004) | | -0.004* (-0.008,-0.001) | | -0.006* (-0.008,-0.004) | -0.006* (-0.008,-0.004) | -0.006* (-0.008,-0.004) | -0.006* (-0.008,-0.005) | -0.007* (-0.008,-0.005) | -0.007* (-0.008,-0.005) | -0.006* (-0.008,-0.005) | -0.005* (-0.007,-0.002) | -0.005* (-0.007,-0.003) | -0.007* (-0.009,-0.005) |

* Indicates statistically significant difference relative to the ERM[08-10] models (trained using 2008-2010) i.e., having a 95% confidence interval that is entirely above or below 0. Abbreviations: DG: domain generalization; UDA: unsupervised domain adaptation; Med: median; CI: confidence interval; LOS: length of stay; ERM: empirical risk minimization; IRM: invariant risk minimization; GroupDRO: group distributionally robust optimization; AL: adversarial learning; CORAL: correlational alignment; AUROC: are under the receiver operator characteristic curve; AUPRC: area under the precision recall curve; ACE: absolute calibration error.

**eTable 2.** Difference in the performance measures of DG, UDA, and ERM[08-16] models relative to ERM[17-19] in 2017–2019.

|  |  | **Med(95%CI)** | **Med Difference (95% CI)** | | | | | | | | | | | | | |
| --- | --- | --- | --- | --- | --- | --- | --- | --- | --- | --- | --- | --- | --- | --- | --- | --- |
|  |  |  | **Domain Generalization** | | | | | | **Unsupervised Domain Adaptation** | | | | | | | |
|  |  |  |  |  |  |  | |  | **100 OOD Samples** | | **500 OOD Samples** | | **1000 OOD Samples** | | **1500 OOD Samples** | |
|  | **Metric** | **ERM**  **[17–19]** | **ERM**  **[08-16]** | **IRM** | **GroupDRO** | | **AL** | **CORAL** | **AL** | **CORAL** | **AL** | **CORAL** | **AL** | **CORAL** | **AL** | **CORAL** |
| **Long LOS** | | |  |  |  |  | |  |  |  |  |  |  |  |  |  |
|  | **AUROC** | ***0.702 (0.696,0.708)*** | -0.024* (-0.030,-0.019) | -0.024* (-0.029,-0.019) | -0.023* (-0.028,-0.017) | | -0.023* (-0.029,-0.018) | -0.024* (-0.029,-0.019) | -0.023* (-0.028,-0.018) | -0.023* (-0.028,-0.018) | -0.024* (-0.029,-0.019) | -0.022* (-0.027,-0.016) | -0.023* (-0.028,-0.018) | -0.022* (-0.027,-0.017) | -0.023* (-0.028,-0.018) | -0.023* (-0.028,-0.018) |
|  | **AUPRC** | ***0.542 (0.532,0.552)*** | -0.009* (-0.016,-0.002) | -0.006 (-0.013,0.000) | -0.008* (-0.015,-0.001) | | -0.007* (-0.014,-0.000) | -0.007* (-0.014,-0.000) | -0.007 (-0.013,0.000) | -0.007* (-0.014,-0.000) | -0.008* (-0.014,-0.001) | -0.006 (-0.013,0.001) | -0.007* (-0.014,-0.000) | -0.005 (-0.012,0.001) | -0.007* (-0.014,-0.000) | -0.008* (-0.014,-0.001) |
|  | **ACE** | ***0.032 (0.029,0.036)*** | 0.029* (0.024,0.033) | 0.022* (0.016,0.027) | 0.044* (0.039,0.049) | | 0.028* (0.022,0.033) | 0.029* (0.023,0.034) | 0.025* (0.020,0.030) | 0.027* (0.022,0.032) | 0.025* (0.020,0.030) | 0.025* (0.020,0.030) | 0.026* (0.021,0.031) | 0.028* (0.023,0.033) | 0.028* (0.023,0.033) | 0.032* (0.027,0.037) |
| **Sepsis** | | |  |  |  |  | |  |  |  |  |  |  |  |  |  |
|  | **AUROC** | ***0.735 (0.724,0.746)*** | -0.041* (-0.049,-0.033) | -0.045* (-0.053,-0.037) | -0.071* (-0.081,-0.062) | | -0.042* (-0.050,-0.034) | -0.044* (-0.053,-0.036) | -0.043* (-0.052,-0.035) | -0.043* (-0.051,-0.034) | -0.044* (-0.053,-0.035) | -0.043* (-0.052,-0.035) | -0.041* (-0.050,-0.033) | -0.044* (-0.052,-0.035) | -0.042* (-0.050,-0.033) | -0.045* (-0.053,-0.036) |
|  | **AUPRC** | ***0.298 (0.280,0.317)*** | -0.104* (-0.118,-0.090) | -0.106* (-0.120,-0.092) | -0.130* (-0.144,-0.116) | | -0.105* (-0.119,-0.092) | -0.106* (-0.121,-0.092) | -0.107* (-0.121,-0.093) | -0.106* (-0.119,-0.092) | -0.105* (-0.119,-0.091) | -0.105* (-0.119,-0.091) | -0.105* (-0.119,-0.091) | -0.106* (-0.120,-0.092) | -0.104* (-0.117,-0.090) | -0.108* (-0.122,-0.094) |
|  | **ACE** | ***0.013 (0.011,0.016)*** | 0.018* (0.014,0.022) | 0.019* (0.015,0.023) | 0.029* (0.025,0.033) | | 0.017* (0.013,0.021) | 0.016* (0.012,0.021) | 0.019* (0.015,0.024) | 0.020* (0.016,0.024) | 0.021* (0.016,0.025) | 0.019* (0.015,0.023) | 0.020* (0.016,0.024) | 0.019* (0.014,0.023) | 0.018* (0.014,0.022) | 0.020* (0.016,0.024) |
| **Mortality** | | |  |  |  |  | |  |  |  |  |  |  |  |  |  |
|  | **AUROC** | ***0.903 (0.897,0.909)*** | -0.008* (-0.012,-0.005) | -0.058* (-0.066,-0.051) | -0.010* (-0.014,-0.006) | | -0.010* (-0.014,-0.006) | -0.010* (-0.014,-0.006) | -0.010* (-0.015,-0.006) | -0.010* (-0.014,-0.006) | -0.009* (-0.013,-0.005) | -0.011* (-0.015,-0.007) | -0.007* (-0.011,-0.003) | -0.010* (-0.014,-0.006) | -0.010* (-0.014,-0.006) | -0.009* (-0.013,-0.005) |
|  | **AUPRC** | ***0.494 (0.472,0.515)*** | 0.049* (0.038,0.061) | 0.005 (-0.010,0.021) | 0.048* (0.036,0.059) | | 0.047* (0.036,0.059) | 0.045* (0.033,0.056) | 0.044* (0.032,0.055) | 0.049* (0.037,0.061) | 0.049* (0.037,0.060) | 0.045* (0.033,0.056) | 0.051* (0.040,0.063) | 0.047* (0.035,0.058) | 0.049* (0.038,0.061) | 0.048* (0.036,0.059) |
|  | **ACE** | ***0.014 (0.012,0.015)*** | 0.003* (0.002,0.004) | 0.030* (0.028,0.032) | 0.004* (0.003,0.005) | | 0.004* (0.003,0.005) | 0.005* (0.003,0.006) | 0.002* (0.001,0.002) | 0.004* (0.003,0.005) | 0.003* (0.002,0.004) | 0.004* (0.003,0.005) | 0.002* (0.001,0.003) | 0.003* (0.002,0.005) | 0.003* (0.002,0.004) | 0.004* (0.003,0.005) |
| **Invasive Ventilation** | | |  |  |  |  | |  |  |  |  |  |  |  |  |  |
|  | **AUROC** | ***0.880 (0.872,0.888)*** | 0.005* (0.001,0.009) | 0.005* (0.000,0.009) | 0.004 (-0.001,0.008) | | 0.004 (-0.001,0.008) | 0.005* (0.001,0.009) | 0.004 (-0.000,0.008) | 0.004 (-0.000,0.009) | 0.003 (-0.001,0.008) | 0.006* (0.002,0.010) | 0.005* (0.000,0.009) | 0.006* (0.002,0.011) | 0.000 (-0.004,0.005) | 0.003 (-0.001,0.007) |
|  | **AUPRC** | ***0.622 (0.601,0.642)*** | 0.025* (0.016,0.035) | 0.024* (0.014,0.033) | 0.017* (0.008,0.027) | | 0.020* (0.010,0.030) | 0.027* (0.018,0.037) | 0.021* (0.012,0.031) | 0.027* (0.017,0.037) | 0.020* (0.011,0.029) | 0.027* (0.018,0.037) | 0.021* (0.011,0.031) | 0.028* (0.019,0.038) | 0.016* (0.006,0.026) | 0.025* (0.016,0.035) |
|  | **ACE** | ***0.020 (0.019,0.022)*** | -0.001 (-0.003,0.001) | -0.001 (-0.003,0.001) | 0.000 (-0.001,0.002) | | -0.002 (-0.004,0.000) | -0.001 (-0.003,0.001) | -0.001 (-0.003,0.001) | -0.001 (-0.004,0.001) | -0.002 (-0.004,0.000) | -0.002* (-0.004,-0.000) | -0.002 (-0.004,0.000) | 0.000 (-0.002,0.003) | -0.001 (-0.003,0.002) | -0.002* (-0.004,-0.000) |

* Indicates statistically significant difference relative to ERM models (trained using 2017-2019) i.e., having a 95% confidence interval that is entirely above or below 0.

Abbreviations: DG: domain generalization; UDA: unsupervised domain adaptation; Med: median; CI: confidence interval; LOS: length of stay; ERM: empirical risk minimization; IRM: invariant risk minimization; GroupDRO: group distributionally robust optimization; AL: adversarial learning; CORAL: correlational alignment; AUROC: are under the receiver operator characteristic curve; AUPRC: area under the precision recall curve; ACE: absolute calibration error.

**eTable 3.** Difference in the performance measures of DG and UDA models relative to the ERM[08-16] models in 2017-2019.

|  |  | **Med (95% CI)** |  |  | **Med Difference (95% CI)** | | | | | | | | | | |
| --- | --- | --- | --- | --- | --- | --- | --- | --- | --- | --- | --- | --- | --- | --- | --- |
|  |  |  | **Domain Generalization** | | | | **Unsupervised Domain Adaptation** | | | | | | | | |
|  |  |  |  |  |  |  | **100 Unlabeled Samples** | | | **500 Unlabeled Samples** | | **1000 Unlabeled Samples** | | **1500 Unlabeled Samples** | |
|  | **Metric** | **ERM[08-16]** | **IRM** | **GroupDRO** | **AL** | **CORAL** | **AL** | **CORAL** | | **AL** | **CORAL** | **AL** | **CORAL** | **AL** | **CORAL** |
| **Long LOS** | |  |  |  |  |  |  |  |  | |  |  |  |  |  |
|  | **AUROC** | ***0.678 (0.671,0.684)*** | 0.000 (-0.002,0.002) | 0.002 (-0.001,0.004) | 0.001 (-0.001,0.003) | 0.000 (-0.002,0.002) | 0.001 (-0.001,0.003) | 0.001 (-0.001,0.003) | | 0.000 (-0.002,0.003) | 0.003* (0.000,0.005) | 0.001 (-0.001,0.004) | 0.002 (-0.000,0.005) | 0.001 (-0.001,0.003) | 0.001 (-0.001,0.004) |
|  | **AUPRC** | ***0.533 (0.522,0.543)*** | 0.003* (0.000,0.006) | 0.001 (-0.002,0.005) | 0.002 (-0.001,0.005) | 0.002 (-0.001,0.005) | 0.002 (-0.001,0.006) | 0.002 (-0.002,0.006) | | 0.001 (-0.002,0.005) | 0.003 (-0.000,0.007) | 0.002 (-0.002,0.006) | 0.004 (-0.000,0.008) | 0.002 (-0.002,0.006) | 0.002 (-0.002,0.005) |
|  | **ACE** | ***0.061 (0.057,0.065)*** | -0.007* (-0.008,-0.006) | 0.016* (0.014,0.017) | -0.001 (-0.002,0.000) | 0.000 (-0.001,0.001) | -0.003* (-0.005,-0.002) | -0.002* (-0.003,-0.000) | | -0.003* (-0.005,-0.002) | -0.004* (-0.005,-0.002) | -0.002* (-0.004,-0.000) | -0.001 (-0.002,0.001) | -0.001 (-0.002,0.001) | 0.003* (0.002,0.005) |
| **Sepsis** | |  |  |  |  |  |  |  | |  |  |  |  |  |  |
|  | **AUROC** | ***0.694 (0.683,0.705)*** | -0.004* (-0.007,-0.001) | -0.030* (-0.036,-0.025) | -0.001 (-0.004,0.002) | -0.003* (-0.006,-0.000) | -0.002 (-0.006,0.002) | -0.002 (-0.005,0.002) | | -0.003 (-0.007,0.001) | -0.002 (-0.006,0.002) | -0.000 (-0.004,0.004) | -0.003 (-0.007,0.001) | -0.001 (-0.004,0.003) | -0.004 (-0.007,0.000) |
|  | **AUPRC** | ***0.195 (0.184,0.206)*** | -0.002 (-0.005,0.002) | -0.026* (-0.031,-0.022) | -0.002 (-0.005,0.001) | -0.003 (-0.006,0.001) | -0.003 (-0.008,0.001) | -0.002 (-0.006,0.002) | | -0.002 (-0.006,0.003) | -0.001 (-0.006,0.003) | -0.001 (-0.006,0.004) | -0.002 (-0.007,0.002) | 0.000 (-0.005,0.005) | -0.004 (-0.008,0.000) |
|  | **ACE** | ***0.031 (0.028,0.034)*** | 0.001 (-0.001,0.002) | 0.011* (0.009,0.013) | -0.001 (-0.002,0.000) | -0.002* (-0.003,-0.000) | 0.001 (-0.000,0.003) | 0.002* (0.000,0.004) | | 0.003* (0.001,0.004) | 0.001 (-0.001,0.002) | 0.002* (0.000,0.003) | 0.001 (-0.001,0.002) | 0.000 (-0.001,0.002) | 0.002* (0.000,0.004) |
| **Mortality** | |  |  |  |  |  |  |  | |  |  |  |  |  |  |
|  | **AUROC** | ***0.894 (0.887,0.901)*** | 0.000 (-0.001,0.002) | -0.050* (-0.055,-0.045) | -0.001 (-0.003,0.001) | -0.001 (-0.003,0.000) | -0.001 (-0.004,0.001) | -0.002 (-0.004,0.001) | | -0.001 (-0.004,0.001) | 0.000 (-0.002,0.002) | -0.002 (-0.004,0.001) | 0.002 (-0.001,0.004) | -0.001 (-0.003,0.001) | -0.001 (-0.003,0.001) |
|  | **AUPRC** | ***0.541 (0.520,0.562)*** | 0.002 (-0.004,0.007) | -0.042* (-0.051,-0.033) | 0.000 (-0.005,0.005) | -0.000 (-0.005,0.005) | -0.003 (-0.010,0.005) | -0.004 (-0.012,0.004) | | 0.001 (-0.006,0.008) | 0.001 (-0.006,0.008) | -0.003 (-0.010,0.004) | 0.004 (-0.003,0.011) | -0.001 (-0.008,0.007) | 0.002 (-0.005,0.009) |
|  | **ACE** | ***0.018 (0.016,0.020)*** | -0.001* (-0.001,-0.000) | 0.026* (0.024,0.027) | 0.000 (-0.001,0.001) | 0.000 (-0.000,0.001) | 0.001 (-0.000,0.002) | -0.002* (-0.003,-0.001) | | 0.000 (-0.000,0.001) | -0.001* (-0.002,-0.000) | -0.000 (-0.001,0.001) | -0.001* (-0.002,-0.001) | -0.000 (-0.001,0.000) | -0.001* (-0.002,-0.000) |
| **Invasive Ventilation** | | |  |  |  |  |  |  | |  |  |  |  |  |  |
|  | **AUROC** | ***0.885 (0.877,0.893)*** | -0.000 (-0.003,0.002) | -0.001 (-0.004,0.001) | -0.001 (-0.003,0.001) | -0.000 (-0.002,0.002) | -0.001 (-0.004,0.002) | -0.001 (-0.003,0.002) | | -0.002 (-0.004,0.001) | 0.001 (-0.002,0.004) | -0.000 (-0.003,0.002) | 0.001 (-0.002,0.005) | -0.005 (-0.009,0.000) | -0.002 (-0.005,0.001) |
|  | **AUPRC** | ***0.647 (0.627,0.666)*** | -0.002 (-0.009,0.006) | -0.008* (-0.015,-0.001) | -0.005 (-0.012,0.001) | 0.002 (-0.004,0.009) | -0.004 (-0.013,0.005) | 0.002 (-0.007,0.011) | | -0.005 (-0.014,0.004) | 0.002 (-0.006,0.011) | -0.004 (-0.014,0.005) | 0.003 (-0.007,0.013) | -0.010 (-0.020,0.000) | 0.000 (-0.008,0.009) |
|  | **ACE** | ***0.019 (0.018,0.020)*** | 0.000 (-0.001,0.001) | 0.002 (-0.000,0.004) | -0.000 (-0.001,0.001) | 0.000 (-0.001,0.001) | -0.000 (-0.001,0.001) | -0.000 (-0.001,0.001) | | -0.001 (-0.001,0.000) | -0.001 (-0.002,0.000) | -0.000 (-0.001,0.001) | 0.001 (-0.001,0.004) | 0.001 (-0.001,0.002) | -0.001 (-0.002,0.001) |

* Indicates statistically significant difference relative to ERM[08-16] models, i.e., having a 95% confidence interval that is entirely above or below 0.

Abbreviations: DG: domain generalization; UDA: unsupervised domain adaptation; Med: median; CI: confidence interval; LOS: length of stay; ERM: empirical risk minimization; IRM: invariant risk minimization; GroupDRO: group distributionally robust optimization; AL: adversarial learning; CORAL: correlational alignment; AUROC: are under the receiver operator characteristic curve; AUPRC: area under the precision recall curve; ACE: absolute calibration error.

**eTable 4.** Performance measures (median (95% CI)) of DG, UDA and ERM[08-16] models in the test sets of 2008–2016.

|  |  | **Domain Generalization** | | | | **Unsupervised Domain Adaptation** | | | | | | | |
| --- | --- | --- | --- | --- | --- | --- | --- | --- | --- | --- | --- | --- | --- |
|  |  |  |  |  |  | **100 OOD Samples** | | **500 OOD Samples** | | **1000 OOD Samples** | | **1500 OOD Samples** | |
| **Metric** | ERM  **[08-16]** | IRM | GroupDRO | AL | CORAL | AL | CORAL | AL | CORAL | AL | CORAL | AL | CORAL |
| **Long LOS** |  |  |  |  |  |  |  |  |  |  |  |  |  |
| **AUROC** | 0.744 (0.741,0.748) | 0.743* (0.740,0.747) | 0.741* (0.738,0.745) | 0.744 (0.740,0.747) | 0.744 (0.740,0.747) | 0.744 (0.741,0.748) | 0.744 (0.740,0.747) | 0.744 (0.740,0.747) | 0.745 (0.741,0.748) | 0.744 (0.741,0.748) | 0.744 (0.740,0.748) | 0.744 (0.740,0.747) | 0.744 (0.740,0.748) |
| **AUPRC** | 0.557 (0.550,0.563) | 0.554* (0.548,0.561) | 0.554* (0.548,0.561) | 0.554* (0.548,0.561) | 0.555 (0.549,0.562) | 0.556 (0.550,0.563) | 0.555 (0.549,0.561) | 0.557 (0.550,0.563) | 0.558 (0.551,0.564) | 0.557 (0.550,0.563) | 0.556 (0.550,0.563) | 0.557 (0.551,0.564) | 0.556 (0.550,0.563) |
| **ACE** | 0.022 (0.021,0.023) | 0.022 (0.021,0.024) | 0.029* (0.028,0.031) | 0.023 (0.022,0.025) | 0.022 (0.021,0.024) | 0.021 (0.020,0.022) | 0.021 (0.020,0.023) | 0.022 (0.021,0.023) | 0.022 (0.021,0.024) | 0.022 (0.021,0.023) | 0.022 (0.021,0.023) | 0.022 (0.021,0.024) | 0.022 (0.020,0.023) |
| **Sepsis** |  |  |  |  |  |  |  |  |  |  |  |  |  |
| **AUROC** | 0.815 (0.810,0.821) | 0.815 (0.810,0.821) | 0.803* (0.797,0.809) | 0.817 (0.811,0.823) | 0.812* (0.806,0.818) | 0.815 (0.809,0.821) | 0.816 (0.810,0.822) | 0.816 (0.810,0.822) | 0.815 (0.809,0.821) | 0.815 (0.809,0.821) | 0.815 (0.810,0.821) | 0.816 (0.810,0.822) | 0.816 (0.810,0.822) |
| **AUPRC** | 0.423 (0.410,0.436) | 0.426 (0.412,0.439) | 0.391* (0.378,0.404) | 0.425 (0.411,0.438) | 0.423 (0.409,0.436) | 0.422 (0.409,0.436) | 0.427 (0.414,0.441) | 0.425 (0.412,0.439) | 0.426 (0.412,0.439) | 0.430* (0.416,0.443) | 0.422 (0.408,0.435) | 0.423 (0.409,0.436) | 0.425 (0.412,0.439) |
| **ACE** | 0.013 (0.011,0.014) | 0.013 (0.012,0.015) | 0.019* (0.017,0.021) | 0.013 (0.011,0.015) | 0.014 (0.012,0.016) | 0.013 (0.012,0.015) | 0.013 (0.012,0.015) | 0.013 (0.011,0.014) | 0.012 (0.011,0.014) | 0.011* (0.010,0.013) | 0.013 (0.011,0.015) | 0.013 (0.011,0.014) | 0.012 (0.011,0.014) |
| **Mortality** |  |  |  |  |  |  |  |  |  |  |  |  |  |
| **AUROC** | 0.883 (0.879,0.887) | 0.883 (0.879,0.887) | 0.852* (0.846,0.857) | 0.883 (0.879,0.887) | 0.883 (0.879,0.888) | 0.883 (0.878,0.887) | 0.883 (0.878,0.887) | 0.882 (0.878,0.887) | 0.883 (0.879,0.887) | 0.883 (0.879,0.887) | 0.884 (0.879,0.888) | 0.883 (0.878,0.887) | 0.883 (0.879,0.887) |
| **AUPRC** | 0.425 (0.412,0.438) | 0.425 (0.412,0.438) | 0.415* (0.402,0.428) | 0.427 (0.414,0.440) | 0.429* (0.416,0.442) | 0.427 (0.414,0.439) | 0.427 (0.414,0.440) | 0.425 (0.412,0.438) | 0.427 (0.414,0.440) | 0.426 (0.414,0.439) | 0.426 (0.413,0.438) | 0.425 (0.412,0.438) | 0.428 (0.415,0.441) |
| **ACE** | 0.016 (0.015,0.018) | 0.015* (0.014,0.017) | 0.044* (0.042,0.045) | 0.016 (0.015,0.018) | 0.017* (0.016,0.018) | 0.017* (0.016,0.018) | 0.015* (0.014,0.016) | 0.017 (0.015,0.018) | 0.017* (0.016,0.018) | 0.016 (0.015,0.017) | 0.016* (0.015,0.017) | 0.016 (0.015,0.017) | 0.017 (0.015,0.018) |
| **Invasive Ventilation** | |  |  |  |  |  |  |  |  |  |  |  |  |
| **AUROC** | 0.844 (0.839,0.850) | 0.844 (0.838,0.849) | 0.844 (0.838,0.849) | 0.845 (0.839,0.850) | 0.845 (0.840,0.851) | 0.844 (0.838,0.849) | 0.844 (0.839,0.850) | 0.844 (0.838,0.849) | 0.845 (0.839,0.850) | 0.845 (0.840,0.851) | 0.844 (0.839,0.850) | 0.832* (0.826,0.838) | 0.845 (0.840,0.851) |
| **AUPRC** | 0.585 (0.572,0.597) | 0.583 (0.571,0.595) | 0.581 (0.569,0.594) | 0.584 (0.572,0.596) | 0.582 (0.570,0.594) | 0.582 (0.570,0.595) | 0.582 (0.569,0.594) | 0.584 (0.572,0.596) | 0.585 (0.573,0.597) | 0.584 (0.572,0.596) | 0.580 (0.567,0.592) | 0.571* (0.559,0.584) | 0.583 (0.570,0.595) |
| **ACE** | 0.018 (0.017,0.019) | 0.018 (0.017,0.020) | 0.025* (0.023,0.027) | 0.019* (0.018,0.020) | 0.016* (0.015,0.017) | 0.019* (0.018,0.020) | 0.018* (0.017,0.019) | 0.019* (0.018,0.020) | 0.019* (0.018,0.020) | 0.018 (0.017,0.020) | 0.016 (0.015,0.017) | 0.019 (0.018,0.020) | 0.018 (0.017,0.019) |

* Indicates statistically significant difference relative to ERM[08-16] (trained using 2008-2016).

Abbreviations: DG: domain generalization; UDA: unsupervised domain adaptation; Med: median; CI: confidence interval; LOS: length of stay; ERM DG: empirical risk minimization domain generalization; IRM: invariant risk minimization; GroupDRO: group distributionally robust optimization; AL: adversarial learning; CORAL: correlational alignment; AUROC: are under the receiver operator characteristic curve; AUPRC: area under the precision recall curve; ACE: absolute calibration error.

**eTable 5.** Threshold-based metrics in the Sepsis prediction task across different thresholds.

|  | **0.05** | **0.1** | **0.15** | **0.2** | **0.25** | **0.3** | **0.35** | **0.4** | **0.45** |
| --- | --- | --- | --- | --- | --- | --- | --- | --- | --- |
| **Sensitivity** | | | | | | | | | |
| ERM (08-10🡪08-10) | 0.87 | 0.65 | 0.56 | 0.48 | 0.41 | 0.37 | 0.33 | 0.3 | 0.27 |
| ERM (08-10🡪17-19) | 0.85 | 0.57 | 0.41 | 0.3 | 0.23 | 0.18 | 0.15 | 0.13 | 0.1 |
| ERM[08-16] | 0.9 | 0.76 | 0.62 | 0.53 | 0.45 | 0.37 | 0.29 | 0.21 | 0.07 |
| AL(1500) | 0.9 | 0.75 | 0.62 | 0.53 | 0.44 | 0.36 | 0.29 | 0.2 | 0.07 |
| ERM[17-19] | 0.91 | 0.61 | 0.43 | 0.33 | 0.25 | 0.2 | 0.15 | 0.1 | 0.06 |
| **Specificity** | | | | | | | | | |
| ERM (08-10🡪08-10) | 0.4 | 0.72 | 0.84 | 0.9 | 0.93 | 0.95 | 0.96 | 0.97 | 0.98 |
| ERM (08-10🡪17-19) | 0.35 | 0.64 | 0.76 | 0.83 | 0.86 | 0.89 | 0.9 | 0.92 | 0.93 |
| ERM[08-16] | 0.42 | 0.68 | 0.79 | 0.85 | 0.89 | 0.92 | 0.95 | 0.97 | 0.99 |
| AL(1500) | 0.42 | 0.68 | 0.79 | 0.85 | 0.89 | 0.92 | 0.95 | 0.97 | 0.99 |
| ERM[17-19] | 0.30 | 0.74 | 0.86 | 0.91 | 0.94 | 0.96 | 0.98 | 0.99 | 0.99 |
| There are 100 consecutive patients admitted to the ICU between 2017-2019. Management will differ depending on whether the risk of sepsis is greater than a threshold (column label) at 4 hours after admission. The table below illustrates the impact of temporal dataset shift, updating the model with more recent data, one domain adaptation strategy and for comparative purposes, the results of the most optimistic model where train and test data came from the most recent period (2017-2019). | | | | | | | | | |
| **False Positives for 89 Patients who did Not Develop Sepsis** |  |  |  |  |  |  |  |  |  |
| ERM (08-10🡪08-10) | 53 | 25 | 14 | 9 | 6 | 4 | 3 | 2 | 2 |
| ERM (08-10🡪17-19) | 58 | 32 | 21 | 15 | 12 | 10 | 9 | 7 | 6 |
| ERM[08-16] | 52 | 29 | 19 | 13 | 10 | 7 | 4 | 3 | 1 |
| AL(1500) | 52 | 28 | 19 | 13 | 9 | 7 | 4 | 3 | 1 |
| ERM[17-19] | 62 | 23 | 13 | 8 | 5 | 3 | 2 | 1 | 1 |
| **False Negatives for 11 Patients that Developed Sepsis** |  |  |  |  |  |  |  |  |  |
| ERM (08-10🡪08-10) | 1 | 4 | 5 | 6 | 6 | 7 | 7 | 8 | 8 |
| ERM (08-10🡪17-19) | 2 | 5 | 6 | 8 | 8 | 9 | 9 | 10 | 10 |
| ERM[08-16] | 1 | 3 | 4 | 5 | 6 | 7 | 8 | 9 | 10 |
| AL(1500) | 1 | 3 | 4 | 5 | 6 | 7 | 8 | 9 | 10 |
| ERM[17-19] | 1 | 4 | 6 | 7 | 8 | 9 | 9 | 10 | 10 |

*Outcome prevalence was estimated based on average sepsis prevalence from 2008 to 2019. The table illustrates the results of initial model development with training and evaluation in the earliest period or 2008-2010, ERM (08-10🡪08-10), which represents performance anticipated by clinicians applying the model to patients admitted 2017-2019 if the model is not updated. ERM (08-10🡪17-19) shows actual performance of that model on their patients, or the impact of temporal dataset shift. In other words, the first two rows under “False Negatives for 11 Patients that Developed Sepsis” illustrate the clinical impact of temporal dataset shift for the task with the most extreme dataset shift, namely sepsis. It shows that for the 11 patients who developed sepsis, the false negative rate increased by 1 or 2 patients. The table also shows the impact of retraining with the more updated data, ERM[08-16], and one approach to mitigate dataset shift, namely AL(1500). Results of AL(1500) was almost identical to ERM[08-16] regardless of the threshold used to define sepsis. For illustrative purposes, it also shows the ERM[17-19] in which training and test sets are both 2017-2019 data.

Abbreviations: ERM: empirical risk minimization; AL(1500): adversarial learning under unsupervised domain adaptation with 1500 unlabeled samples.
